# Dietary Patterns and Cardiovascular Risk in Adult Survivors of Childhood Cancer: Findings From the SCCSS-Nutrition Study

**DOI:** 10.64898/2026.09.22.26363621

**Authors:** Ruijie Li, Raquel Revuelta Iniesta, Alan R. Barker, Angeline Chatelan, Lorenz Leuenberger, Yara Shoman, Ben Spycher, Fabiën N. Belle

## Abstract

**Purpose:** To identify dietary patterns among adult childhood cancer survivors (CCSs), examine their determinants, and assess whether they differ according to cardiovascular disease (CVD) risk profiles.

**Methods:** CCSs from the Swiss Childhood Cancer Survivor Study diagnosed before age 21 years (1976-2005), surviving ≥5 years and aged ≥18 years in 2017, completed a food frequency questionnaire (SCCSS-Nutrition). Factor and hierarchical cluster analyses of 52 food groups identified dietary patterns. We categorised CCSs as having CVD, CVD risk factors (obesity, hypertension, diabetes, high cholesterol), or being CVD risk-free. Multinomial logistic regression assessed associations between dietary patterns and CVD risk profiles.

**Results:** Among 802 CCSs (median age: 35 years; IQR: 29-41; 50% male), we identified five dietary patterns: Mediterranean-inspired diet (22%), Western indulgence diet (38%), Western sweet & fatty diet (26%), restrained indulgence diet (9%), plant-based & alcohol-rich diet (6%). Male sex, younger age, German-speaking region, smoking, and living with others were associated with Western indulgence, Western sweet & fatty, and restrained indulgence diets. Higher Swiss socioeconomic position (Swiss-SEP) and education, and not having children, were associated with the plant-based & alcohol-rich diet, higher Swiss-SEP was also associated with the Western indulgence diet. No associations were observed between dietary patterns and CVD risk profiles.

**Conclusions:** Dietary patterns among adult CCSs were socially patterned and differed according to sociodemographic and lifestyle characteristics, but were not associated with CVD risk profiles.

**Implications for Cancer Survivors:** Dietary counselling and broader lifestyle support should be considered as part of long-term survivorship care for CCSs.

## 1. Introduction

Childhood cancer survivors (CCSs) frequently encounter health complications decades earlier than expected, with cardiovascular disease (CVD) emerging as the predominant cause of morbidity and mortality [1]. Long-term effects of cardiotoxic treatments, such as anthracycline and radiotherapy, contribute to increased risks of CVD [1]. Modifiable lifestyle factors, particularly diet, play a critical role in mitigating CVD risks [2]. Adherence to a healthy dietary pattern, such as the Mediterranean diet, rich in fruits, vegetables, whole grains, and unsaturated fats, is associated with a reduced risk of CVD in CCSs [2]. However, CCSs often have suboptimal dietary behaviours like the general population, including low intake of fruits, vegetables, and whole grains, and high consumption of processed foods rich in saturated fats and added sugars [2]. These unhealthy dietary behaviours may increase their already elevated CVD risks.

The dietary behaviours of CCSs may be influenced by their cancer history, treatment-related side effects, and psychosocial factors. CCSs report altered taste preferences, gastrointestinal discomfort, and emotional stress, all of which can shape food choices [3]. Additionally, factors such as socioeconomic status, lifestyle habits, and specific treatment exposures may further influence dietary patterns in CCSs [4]. Analysing overall dietary patterns provides a more comprehensive understanding of diet– disease relationships compared to studies focusing on individual nutrients or food items [5]. Although dietary behaviours in CCSs have been studied, only the St. Jude Lifetime Cohort (SJLIFE) in the U.S. investigated dietary patterns [4].

A better understanding of dietary patterns and their determinants can identify vulnerable groups and may support the development of nutritional interventions to improve long-term health. Therefore, this study aims to: 1) evaluate whether dietary patterns are associated with CVD and CVD risk factors; 2) identify dietary patterns of Swiss CCSs; and 3) assess their sociodemographic and lifestyle determinants.

## 2. Methods

### 2.1. Study Populations

The Swiss Childhood Cancer Survivor Study (SCCSS, www.swiss-ccss.ch) is a population-based, long-term follow-up study of all childhood cancer patients registered in the Swiss Childhood Cancer Registry (ChCR, www.chilhoodcancerregistry.ch). Eligible participants include those diagnosed in Switzerland with leukaemia, lymphoma, central nervous system tumours, malignant solid tumours, or Langerhans cell histiocytosis before age 21, who survived at least five years post-diagnosis, and were alive at study enrolment [6]. The SCCSS is registered at ClinicalTrials.gov (identifier: NCT03297034). Ethical approval was granted by the ethics committee of the canton of Bern, Switzerland (KEK-BE: 166/2014 and 2021-01462).

CCSs were eligible to participate in the SCCSS-Nutrition study if they were diagnosed with childhood cancer between 1976 and 2005, completed a baseline SCCSS questionnaire (2007-2013), and were ≥18 years old at the 2017 follow-up [6]. All eligible CCSs enrolled in SCCSS-Nutrition received a follow-up questionnaire that included a food frequency questionnaire (FFQ). Complete details of the SCCSS-Nutrition study design are available in our previously published protocol [6].

We followed the Strengthening the Reporting of Observational Studies in Epidemiology checklist when we reported this manuscript [7]. We excluded participants from this cross-sectional if they were pregnant or breastfeeding, if dietary data were incomplete, or if total energy intake was implausible (<850 or >4500 kcal/day), based on previously suggested cut-offs for unreliable reporting in this FFQ [8].

### 2.2. Measurements

#### 2.2.1. Food Frequency Questionnaire

At follow-up, we assessed dietary intake of CCSs using a self-administered, semi-quantitative FFQ, which was initially developed and validated for the French-speaking Swiss adult population through comparisons with 24-hour dietary recalls [9–11]. The FFQ captures consumption frequency and portion sizes for 112 fresh and prepared food items over the previous four weeks. Consumption frequencies range from “never in the last four weeks” to “two or more times per day.” Portion sizes were categorised as smaller than, equal to, or larger than a reference size, ranging from the first to the third quartile of portion sizes identified in the 24-hour dietary recalls [11]. We converted reported frequencies and portion sizes into daily intakes (g/day), aggregated them into 52 predefined food groups based on common food categories (Supplementary Table 1), and energy-adjusted (g/1,000 kcal) them.

#### 2.2.2. Sociodemographic, Lifestyle, and Clinical Characteristics

We obtained demographic and lifestyle data from the ChCR and the follow-up questionnaire, including sex, age at survey, language region within Switzerland (German, French or Italian speaking region), migration background (Swiss or having a migration background), educational level (university level or lower), marital status (married or single, widowed or divorced), having children (yes/no), living situation (living alone or with others), smoking status (never, former or current), self-reported weight (without clothing) and height (without shoes). Participants were classified as having a migration background if they were not Swiss citizens at birth, were born outside Switzerland, or had at least one parent without Swiss citizenship.

We calculated body mass index (BMI*, [weight]/[height²],* kg/m²). BMI categories included underweight (<18.5 kg/m²), normal weight (18.5–24.9 kg/m²), overweight (25–29.9 kg/m²), and obesity (≥30 kg/m²) [12].

Physical activity (PA) was assessed with questionnaire items on the frequency and duration of moderate- and higher-intensity activity. Based on WHO guidelines, participants were classified as inactive (<150 min/week) or active (≥150 min/week of moderate-intensity activity, ≥75 min/week of vigorous-intensity activity, or a combination of moderate and vigorous-intensity PA per week) [13].

We used the Swiss neighbourhood index of socioeconomic position (Swiss-SEP) as a validated area-based measure of socioeconomic status [14]. The Swiss-SEP ranges from 0 to 100; a higher value indicates a neighbourhood holds a higher socioeconomic position. We assigned the nearest Swiss-SEP value to the geographical coordinates of the participant’s home address. We classified Swiss-SEP values into tertiles based on the national Swiss population distribution (low, medium, high).

The 36-Item Short Form Survey (SF-36) assessed participants’ health-related quality of life (HRQoL) across eight domains: physical functioning (limitations in daily physical activities); role physical (impact of physical health on work or daily activities); bodily pain (intensity and effect of pain on normal activities); general health (overall perception of personal health); vitality (feelings of energy and fatigue); social functioning (impact of health on social activities); role emotional (impact of emotional health on daily functioning); and mental health (psychological wellbeing including anxiety and depression) [15]. Each domain was scored on a scale from 0 to 100, following standard procedures; higher scores indicated better health status. The SF-36 survey also provided two summary scores: the physical component summary and the mental component summary, each derived from its domain.

We extracted clinical information from the ChCR, including cancer diagnosis, age at diagnosis, time since diagnosis, and treatments. Diagnoses were classified according to the International Classification of Childhood Cancer, 3rd Edition [16]. We categorised radiotherapy as any, cranial (defined as direct brain/skull irradiation), chest (including total body irradiation, mantle-field radiation, or thorax/mediastinal/thoracic spinal irradiation), total body/abdominal radiation, or no radiotherapy. We obtained cumulative radiation doses from medical records and applied Children’s Oncology Group Long-Term Follow-Up classification criteria: cranial (<18 vs ≥18 Gy), chest (<30 vs ≥30 Gy), and total body/abdominal (present vs absent) [17]. Other treatments included glucocorticoids (with prednisone/dexamethasone doses estimated from protocols [18]), anthracyclines, alkylating agents, and haematopoietic stem cell transplantation. We also retrieved records on relapse during follow-up visits.

#### 2.2.3. Cardiovascular Risk Profiles

Assessments of cardiovascular health were based on self-reported data from the SCCSS follow-up questionnaire, completed at the same time as the FFQ. Assessment of cardiovascular conditions was based on answers to closed questions and an additional open option; all medication use was recorded. We classified CCSs into one of three CVD profiles: 1) “CVD,” including suffering from a heart attack, cardiomyopathy, angina pectoris, atrial fibrillation, arteriosclerosis, stroke, transient ischemic attack, and/or deep venous thrombosis; 2) “CVD risk factors,” including having hypertension (repeated high blood pressure measurements or using antihypertensive medication treatment), obesity, diabetes mellitus treated with glucose-lowering medications (e.g. oral agents and/or insulin), and/or high cholesterol defined as treatment with lipid-lowering medications; 3) “CVD risk-free,” if survivors did not report any of the previously mentioned conditions.

### 2.3. Statistical Analyses

To identify dietary patterns, we used a two-step analytical approach. First, we applied factor analysis to the 52 predefined food groups (g/1,000 kcal) to identify foods commonly consumed together (Supplementary Table 1). We based our selection of the number of factors on inspection of the scree plot, eigenvalues, and interpretable dietary patterns. We used varimax rotation to select a 10-factor solution. We calculated factor scores and then used them for cluster analysis. Second, we performed hierarchical cluster analysis with Ward’s linkage method and Euclidean distance to group participants based on their factor scores. We used dendrogram inspection and the pseudo-F statistic to determine the optimal number of clusters (five). Then we used the Mediterranean-inspired pattern as the reference category because its cardioprotective association is well-established.

We evaluated the adherence of each dietary pattern to the dietary reference values established by the European Food Safety Authority (EFSA) [19]. For each nutrient, we calculated individual adherence as the ratio of observed intake to the age- and sex-specific population reference intake (PRI), or to the adequate intake (AI) when PRI was unavailable, multiplied by 100. Pattern-level adherence was expressed as the mean percentage of the reference value achieved. A value of 100% indicated that the recommendation was met; values below 100% indicated intakes fell below the reference value; values above 100% indicated intakes exceeded the reference value.

We used descriptive statistics to summarize sociodemographic, lifestyle factors, and clinical characteristics. We presented continuous variables as medians with interquartile ranges (IQR) or mean with standard deviation (SD), and categorical variables as frequencies and percentages. We compared sociodemographic, lifestyle, and clinical characteristics across diet patterns using the chi-square test for categorical variables and Kruskal-Wallis for continuous variables.

We used multinomial logistic regression to assess associations of dietary patterns with sociodemographic and lifestyle factors by estimating crude and adjusted odds ratios (OR) with 95% confidence intervals (CI). We adjusted for sex, age at survey, and cancer diagnosis, identified a priori in multivariable models.

We used multinomial logistic regression to assess the associations between dietary patterns and CVD risk profiles. Covariates were selected based on prior evidence and results from univariable analyses. We defined two multivariable models: Model 1 was adjusted for sex and age at survey; Model 2 was adjusted for sex, age at survey, Swiss-SEP, smoking status, cancer diagnosis, cranial radiation, and SF-36 physical and mental component summary scores. We used STATA version 18.0 for all analyses (Stata-Corp. 2023; Stata statistical software StataCorp LLC, College Station, Texas, US).

## 3. Results

### 3.1. Study Population

We contacted 1,599 individuals who were alive and with valid address at the time of study, out of 1,749 eligible CCSs (Supplementary Figure 1). Of these, 919 (57% of those contacted) completed and returned the FFQ. We excluded 117 participants: 11 (<1%) who were pregnant or breastfeeding, 35 (2%) with incomplete dietary reporting, and 71 (4%) who reported unreliable energy intake. A total of 802 (50%) CCSs with complete valid FFQ data were included in this study. Participants had a median age of 35 years (IQR: 29-41), and 50% were female (Table 1). The most common cancer diagnoses were leukaemias (31%), lymphomas (22%), and central nervous system tumours (10%), with a median age at diagnosis of 10 years (IQR: 4-14). Included and excluded participants had largely comparable sociodemographic and clinical characteristics (Supplementary Table 2), but excluded participants were more often physically inactive (36% vs. 21%, *p* < 0.001), had higher prevalence of obesity (17% vs. 9%, *p* = 0.040), and were more often diagnosed with central nervous system tumours (20% vs. 10%, *p* = 0.046).

**Table 1.**
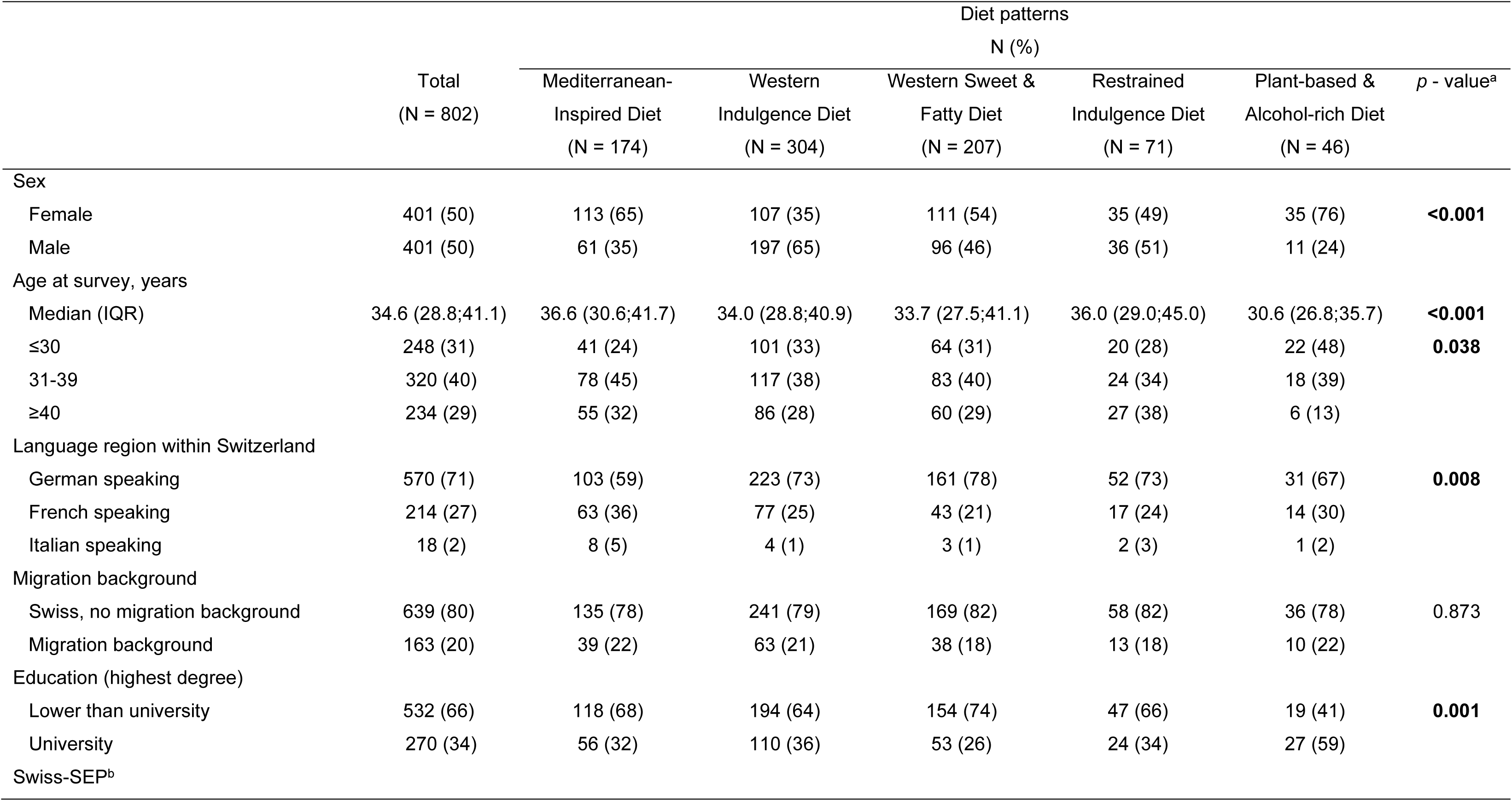
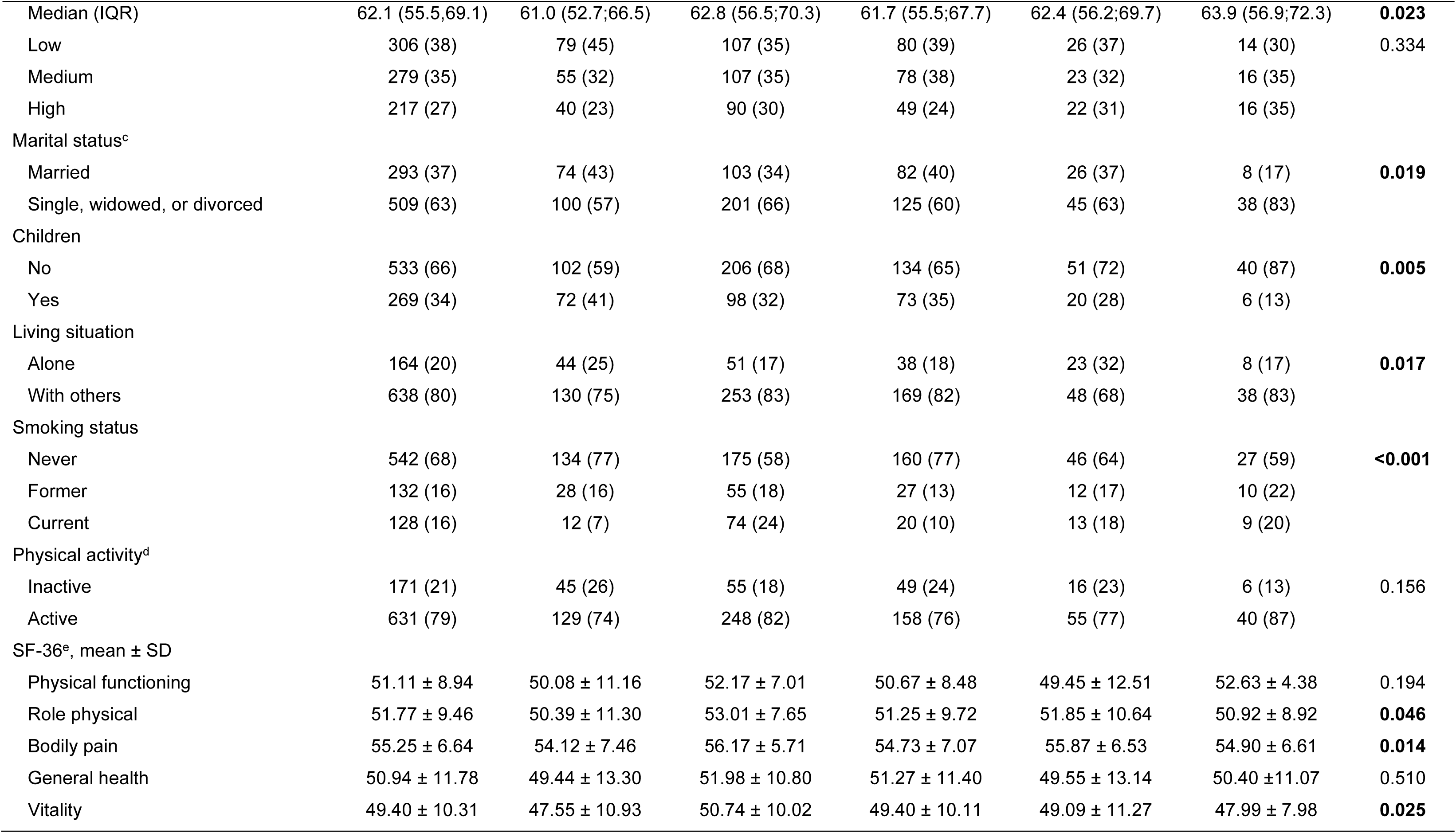
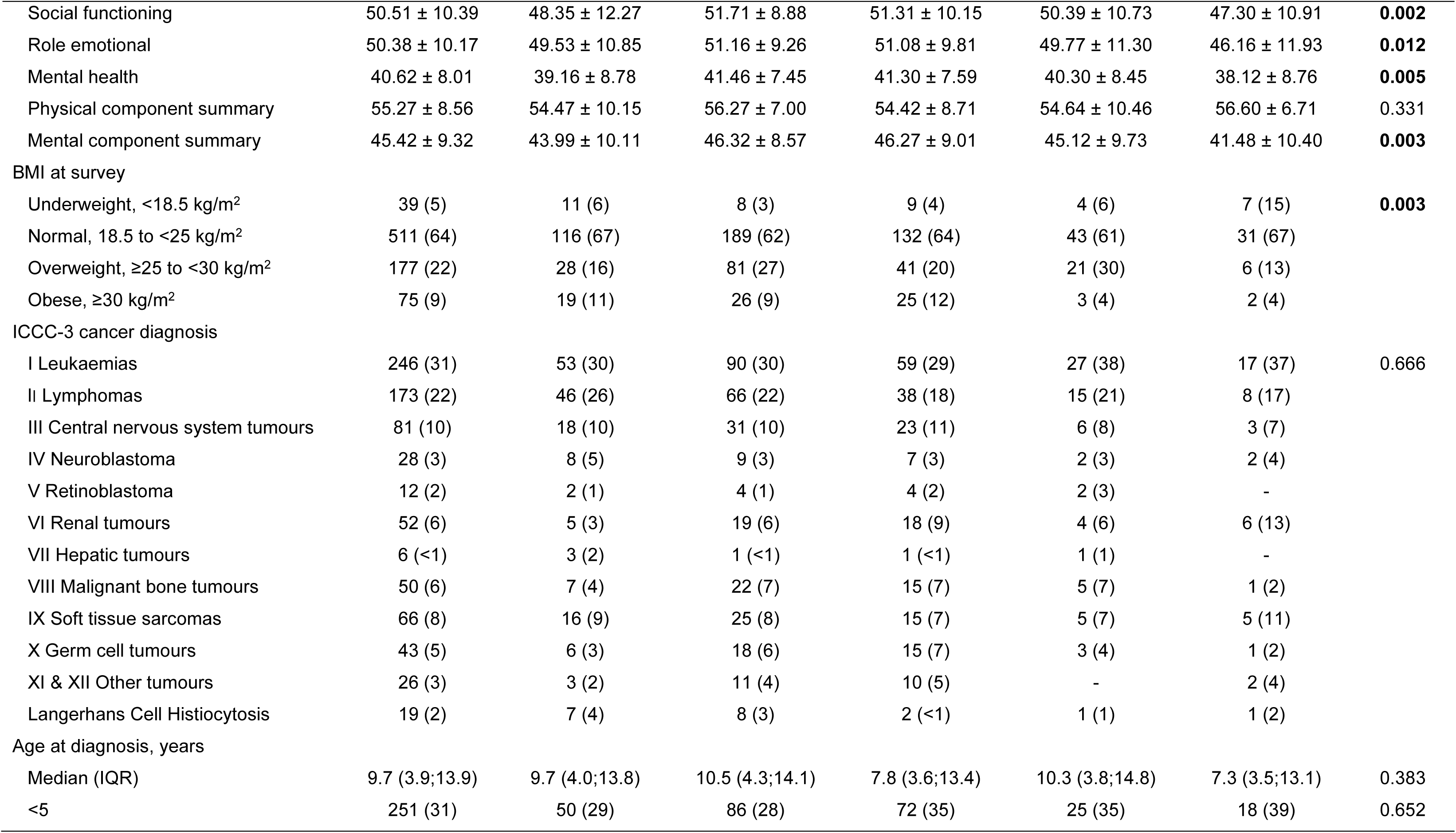
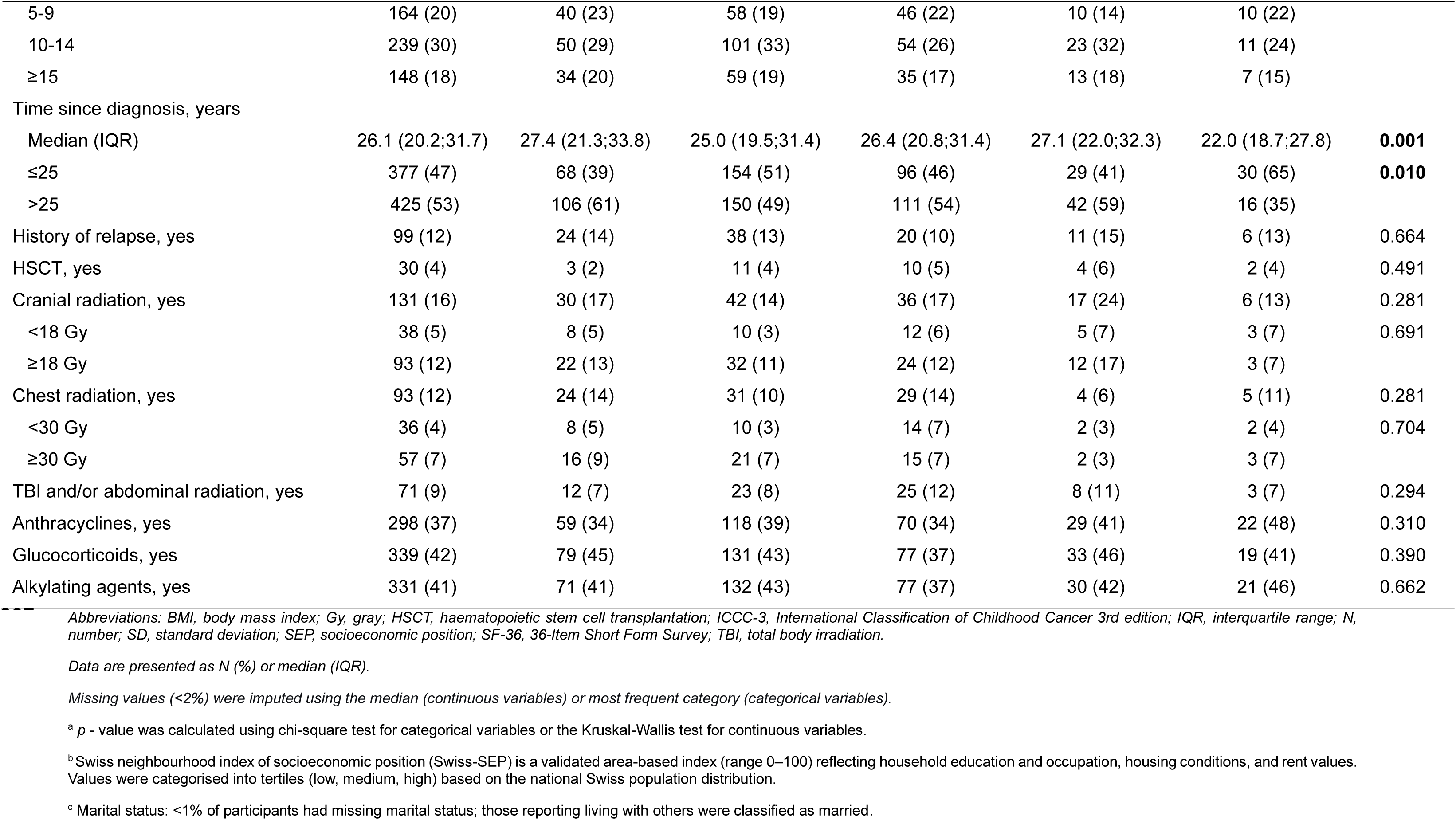
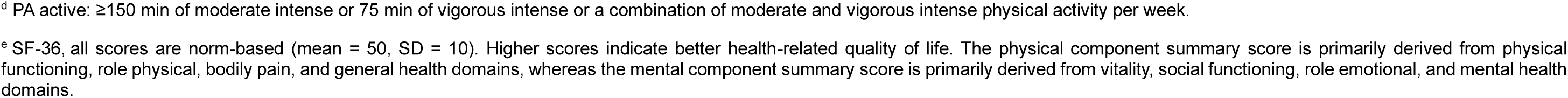
Sociodemographic, lifestyle and clinical characteristics of Swiss adult childhood cancer survivors by dietary patterns.

### 3.2. Dietary Patterns

We identified five distinct dietary patterns, which we label according to the characteristics of the dominant food group (Figure 1, Supplementary Tables 3, 4): 1) *Mediterranean-inspired diet* (N = 174, 22%), featuring abundant fish and shellfish, sweet spreads, fresh fruits and vegetables (green leafy, root, and fruiting vegetables) while low intake of processed/red meats, high-fat dairy, and alcohol; 2) *Western indulgence diet* (N = 304, 38%), with high consumption of fast food (e.g., French fries, pizza, snacks), processed/red meats, and alcohol; 3) *Western sweet & fatty diet* (N = 207, 26%), dominated by high-fat dairy, sugary foods and refined grains; 4) *restrained indulgence diet* (N = 71, 9%), with high intake of processed/red meats, high-fat dairy, and sugar and sweeteners, a low intake of fruits and vegetables, fast food, sweet spreads, and alcohol; and 5) *plant-based & alcohol-rich diet* (N = 46, 6%), marked by high fresh fruits and vegetables, and alcohol intake, but minimal processed/red meat and seafood. Sex distribution differed across diet patterns (p < 0.001, Table 1). Women were more likely to follow the plant-based & alcohol-rich (76%) or Mediterranean-inspired diet (65%), while men were more likely to follow a Western indulgence diet (65%). Those who followed the Mediterranean-inspired diet adhered most closely to EFSA recommendations (Figure 2) for balanced macronutrients, high fibre, and favourable calcium, iron, and potassium, though vitamin D in that group remained low.

**Figure 1.**
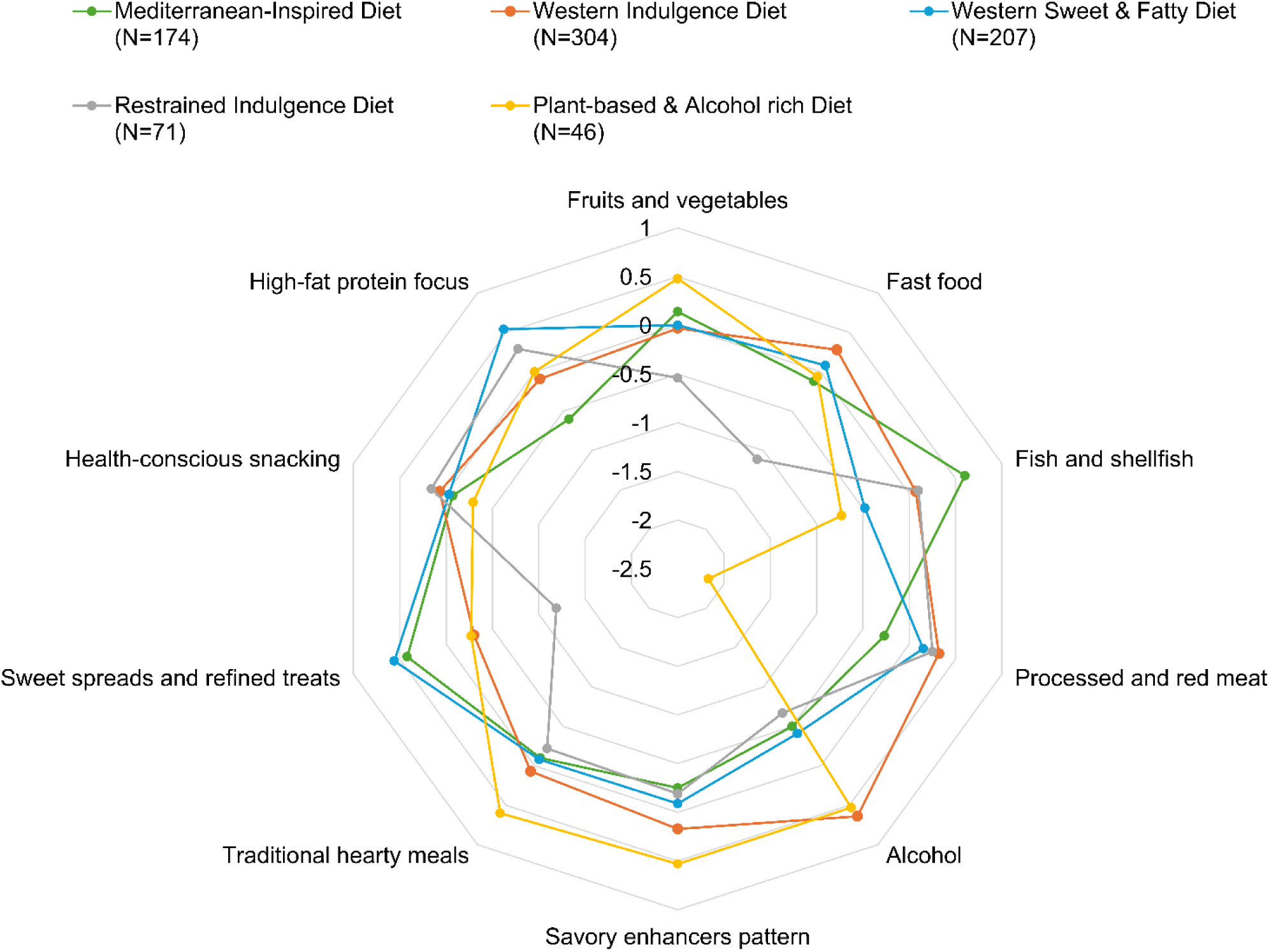
Radar plot of selected factors based on food group intake by identified dietary patterns in Swiss childhood cancer survivors (N=802). The original 52 food groups were reduced to 10 factors through factor analysis. The radar chart displays standardised mean intakes (z-scores) of the 10 factors across five dietary patterns.

**Figure 2.**
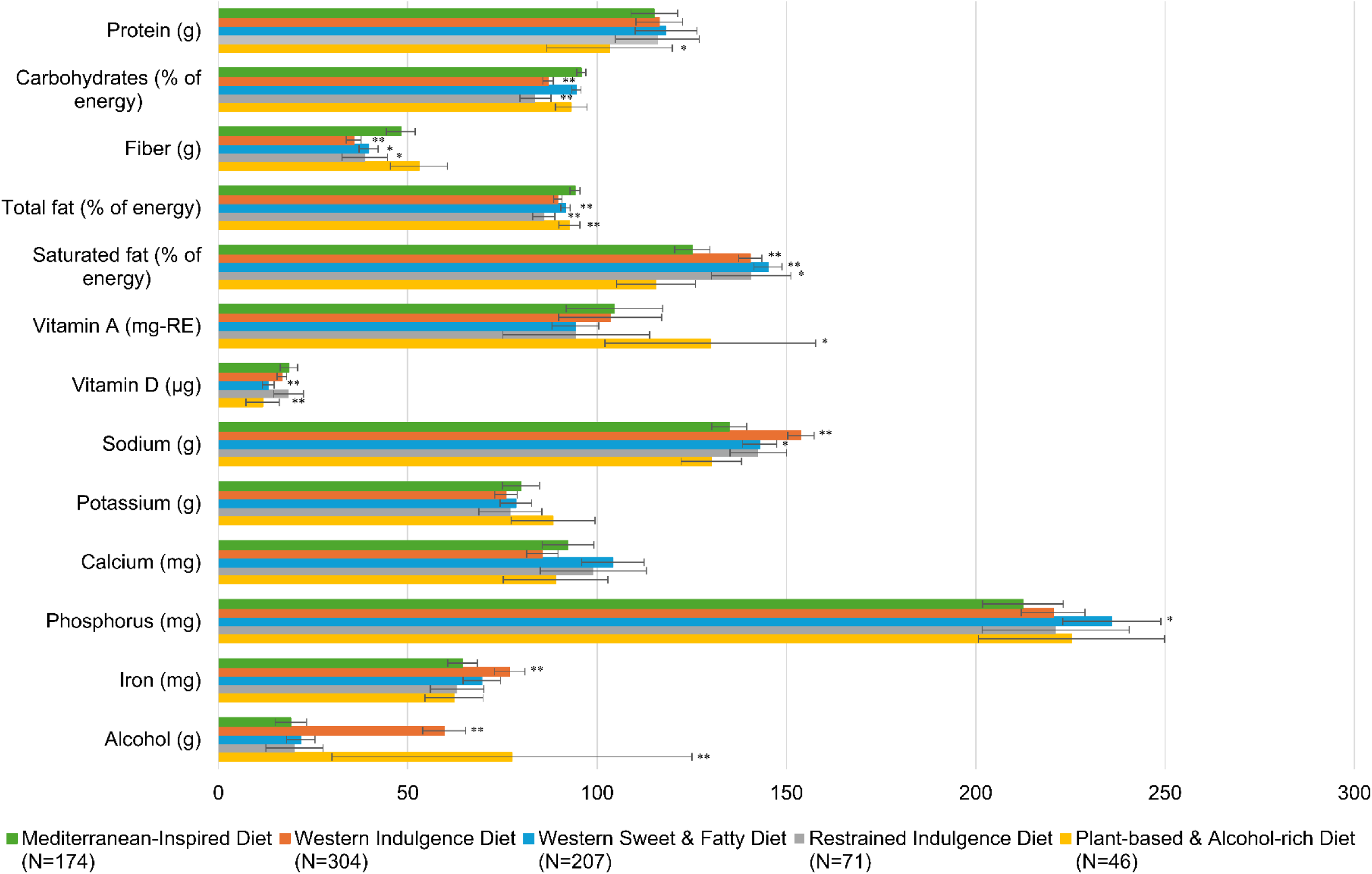
Nutrient intakes in Swiss childhood cancer survivors (N=802) are expressed as the mean percentage of the age- and sex-specific EFSA population reference intake (PRI/AI) achieved (individual intake/ reference intake × 100) across five dietary patterns. Bars represent means with 95% CIs. For alcohol, the maximum recommended limit was used. A value of 100% indicates that the recommendation was met. *p < 0.05; **p < 0.001 vs. the Mediterranean-inspired diet (Mann–Whitney test).

### 3.3. Determinants of Dietary Patterns

Univariable regression results are presented in Supplementary Table 5. Male CCSs were more likely to follow non-Mediterranean diet patterns, particularly the Western indulgence diet. Younger CCSs more likely to follow plant-based & alcohol-rich, Western sweet & fatty, and Western indulgence diets (Table 2, multivariable models). Conversely, those residing in French- and Italian-speaking regions were less likely to follow the Western sweet & fatty or Western indulgence diets. Higher educational attainment was associated with the plant-based & alcohol-rich diet, whereas having children reduced the likelihood of this pattern. Higher Swiss-SEP was associated with both the plant-based & alcohol-rich and Western indulgence diets. Living with others increased the odds of following the Western indulgence diet. Current smokers more often followed the restrained indulgence, plant-based & alcohol-rich, and Western indulgence diets. We found no meaningful evidence of associations between dietary patterns and HRQoL indicators (SF-36), although some reached statistical significance (p < 0.05, ORs ≈ 1.0).

**Table 2.**
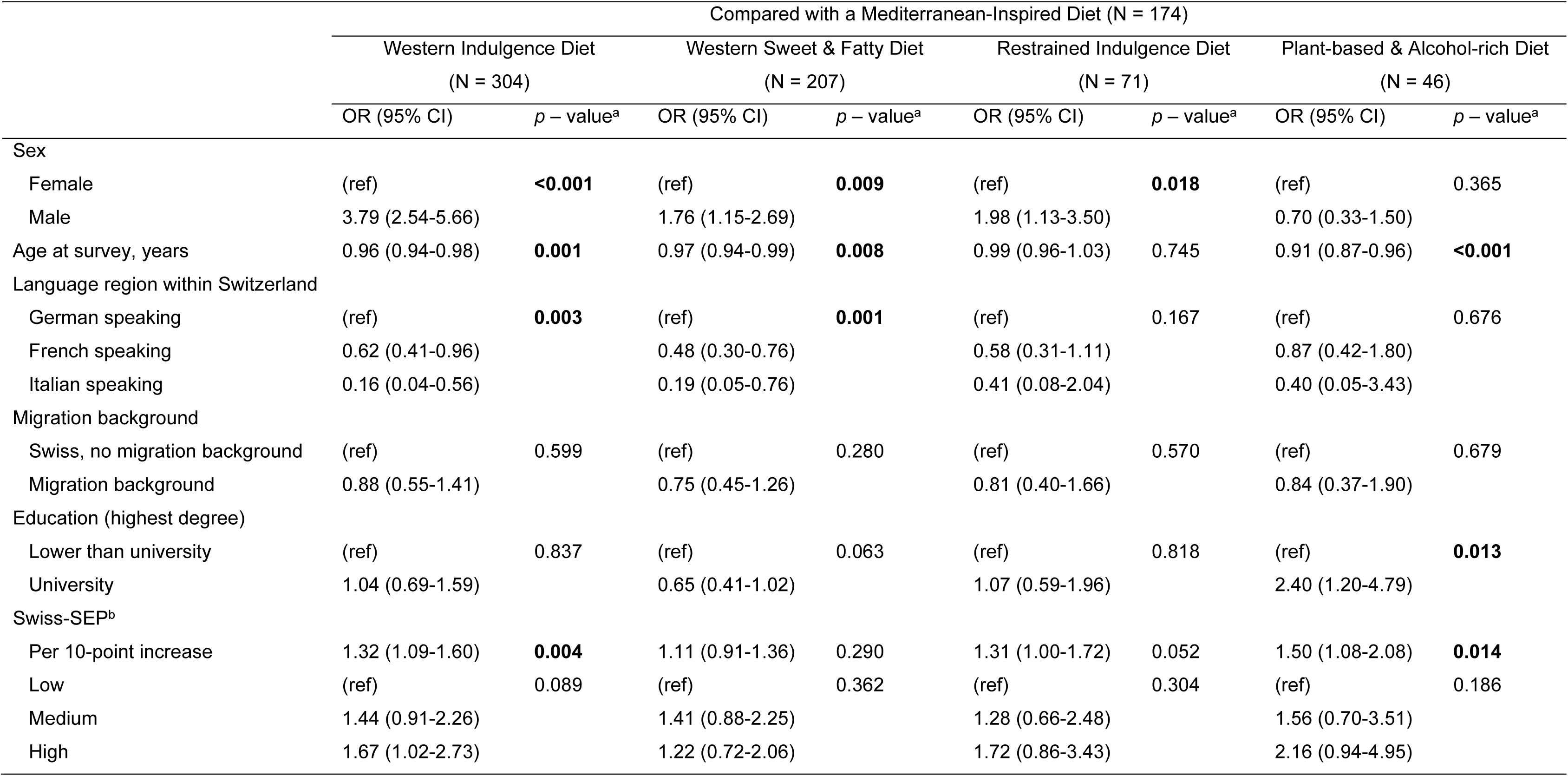
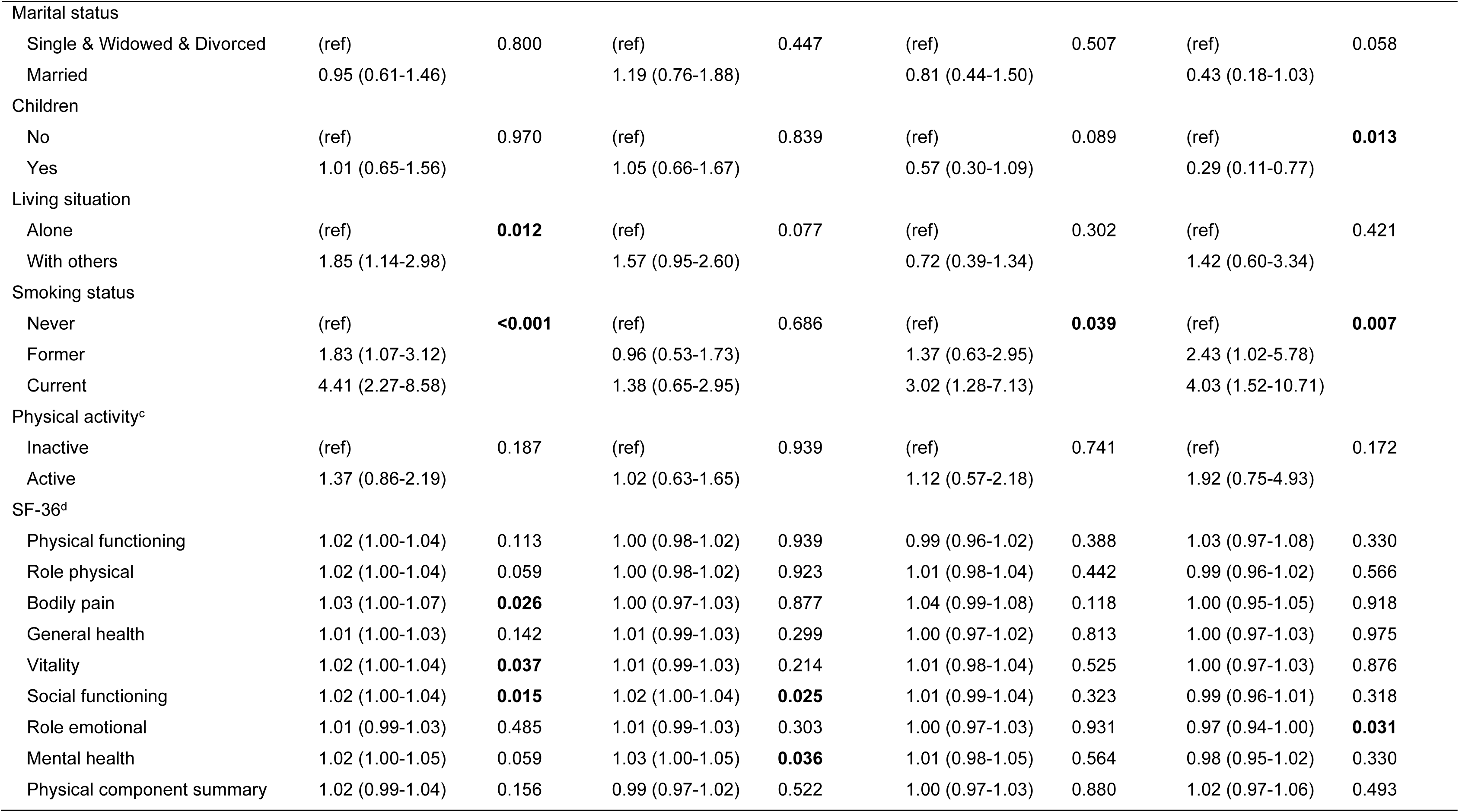
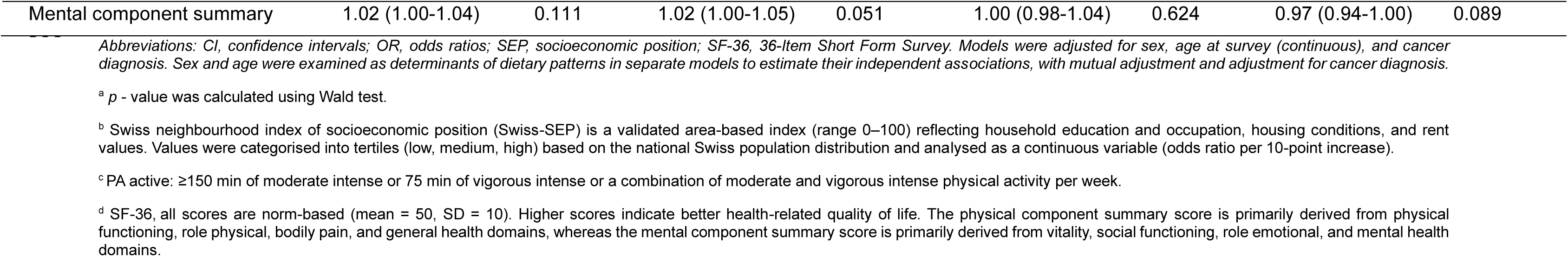
Socio-demographic and lifestyle determinants for dietary patterns in adult Swiss childhood cancer survivors (from multivariable multinomial logistic analysis, adjusted for sex, age at survey, cancer diagnosis).

### 3.4. Dietary Patterns and Cardiovascular Health

Among CCSs, 14% (N = 114) reported at least one CVD, 11% (N = 88) reported at least one CVD risk factor (obesity, hypertension, diabetes, high cholesterol), and 75% (N = 600) were reported CVD risk-free. The prevalence of CVD and CVD risk factors was similar across dietary patterns (p = 0.728; Figure 3).

**Figure 3.**
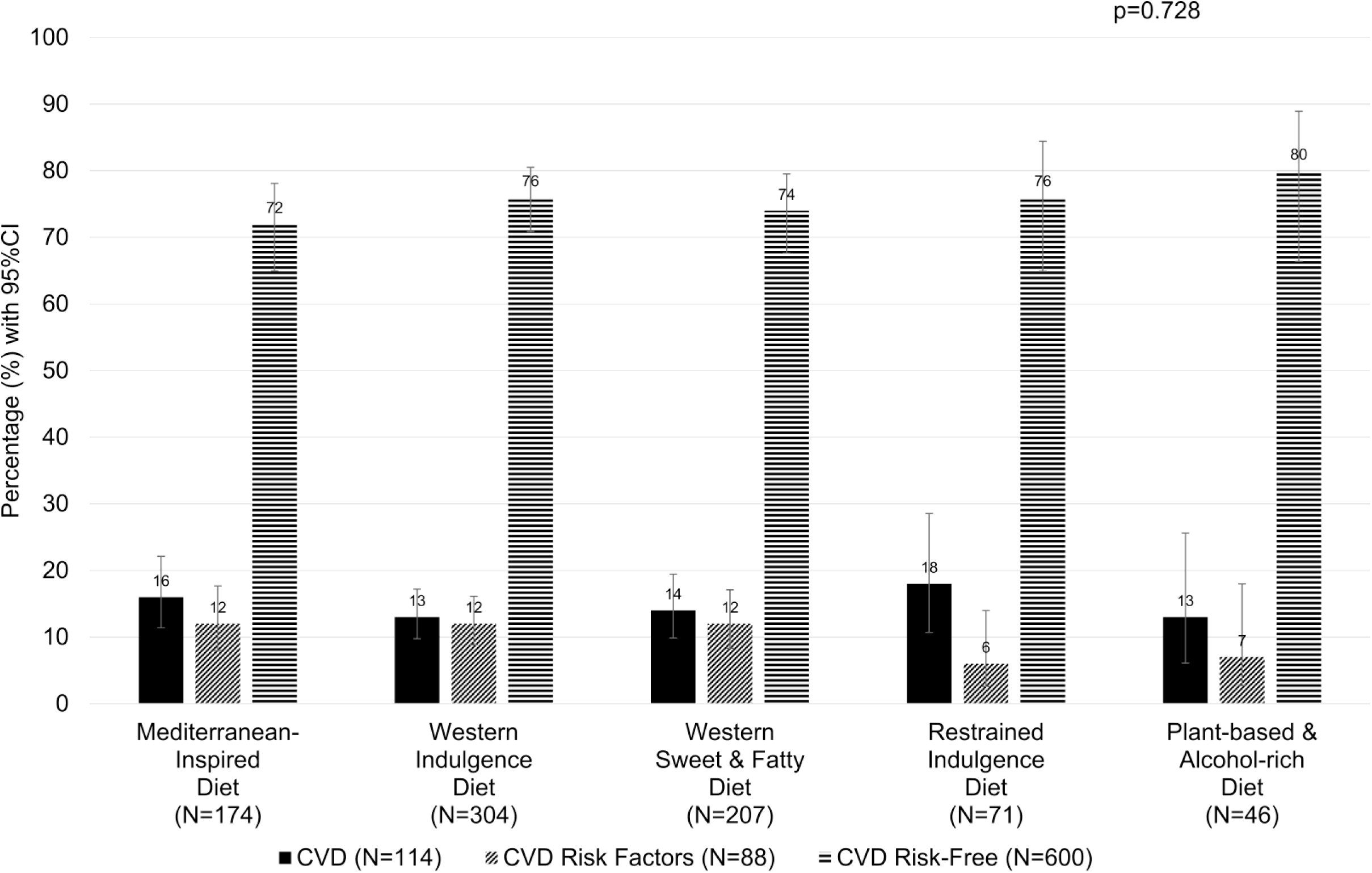
Distribution of CVD risk profiles across diet patterns in Swiss adult childhood cancer survivors. 75% CCSs (N=600) are CVD risk-free, meaning that CCSs did not report any CVD or CVD risk factor. 11% CCSs (N=88) had at least one CVD risk factor, of whom 75 (40%) had obesity (BMI ≥30 kg/m²), 21 (11%) had repeated high blood pressure, 13 (7%) had high cholesterol, and 8 (4%) had diabetes. 14% CCSs (N=114) had CVD, of whom 69 (61%) CCSs had an atrial fibrillation, 21 (18%) deep venous thrombosis, 20 (18%) cardiomyopathy, 10 (9%) stroke/transient ischemic attack, 8 (7%) angina pectoris, 5 (4%) a heart attack, and 5 (4%) arteriosclerosis. Error bars represent 95% CI for prevalence estimates. Differences across dietary patterns were assessed using the chi-square test.

Univariable associations between sociodemographic, lifestyle, and clinical characteristics and CVD risk profiles are shown in Supplementary Table 6. Compared with the Mediterranean-inspired diet, we found no statistically significant associations between other dietary patterns and CVD or CVD risk factors in crude or adjusted models (Table 3).

**Table 3.** Association of dietary patterns with CVD risk profiles in Swiss adult childhood cancer survivors: multivariable multinomial logistic regression results.

| Compared to Mediterranean-Inspired Diet patterns | Crude |  | Model 1<sup>a</sup> |  | Model 2<sup>b</sup> |  |
| --- | --- | --- | --- | --- | --- | --- |
|  | OR (95% CI) | *p* - value<sup>c</sup> | OR (95% CI) | *p* - value<sup>c</sup> | OR (95% CI) | *p* - value<sup>c</sup> |
| CVD<sup>d</sup> vs. CVD risk-free<sup>e</sup> |  |  |  |  |  |  |
| Western Indulgence Diet | 0.77 (0.45-1.32) | 0.344 | 0.97 (0.55-1.71) | 0.913 | 1.02 (0.57-1.85) | 0.941 |
| Western Sweet & Fatty Diet | 0.92 (0.52-1.62) | 0.760 | 1.09 (0.61-1.96) | 0.775 | 1.02 (0.57-1.85) | 0.858 |
| Restrained Indulgence Diet | 1.12 (0.54-2.34) | 0.756 | 1.17 (0.55-2.50) | 0.671 | 1.16 (0.53-2.53) | 0.858 |
| Plant-based & Alcohol-rich Diet | 0.76 (0.29-1.97) | 0.568 | 1.03 (0.39-2.75) | 0.952 | 1.02 (0.57-1.85) | 0.941 |
| CVD risk factors<sup>f</sup> vs. CVD risk-free |  |  |  |  |  |  |
| Western Indulgence Diet | 0.94 (0.53-1.68) | 0.832 | 1.02 (0.56-1.88) | 0.948 | 1.14 (0.61-2.15) | 0.685 |
| Western Sweet & Fatty Diet | 0.94 (0.50-1.77) | 0.851 | 1.06 (0.55-2.02) | 0.865 | 1.06 (0.54-2.05) | 0.874 |
| Restrained Indulgence Diet | 0.44 (0.15-1.36) | 0.154 | 0.43 (0.14-1.33) | 0.144 | 0.42 (0.13-1.31) | 0.135 |
| Plant-based & Alcohol-rich Diet | 0.49 (0.14-1.72) | 0.264 | 0.70 (0.19-2.52) | 0.585 | 0.71 (0.19-2.62) | 0.610 |

Abbreviations: CVD, cardiovascular disease; CI, confidence intervals; OR, odds ratios.
<sup>a</sup> Model 1: Adjusted for sex, age at survey (continuous).
<sup>b</sup> Model 2: Adjusted for sex, age at survey (continuous), Swiss-SEP, smoking status, cancer diagnosis, cranial radiation, SF-36: physical and mental component summary scores.
<sup>c</sup> *p* - value was calculated using Wald test.
<sup>d</sup> "CVD" including atrial fibrillation, deep venous thrombosis, cardiomyopathy, stroke/transient ischemic attack, angina pectoris, heart attack, arteriosclerosis.
<sup>e</sup> "CVD risk-free" if survivors did not report any CVD conditions and CVD risk factors.
<sup>f</sup> "CVD risk factors" including hypertension (repeated high blood pressure measurements or antihypertensive medication treatment), obesity, diabetes mellitus treated with either tablets or insulin, and/or high cholesterol defined as treatment with lipid-lowering medications.

## 4. Discussion

We identified five distinct dietary patterns in this nationwide study of Swiss adult CCSs: Mediterranean-inspired, Western indulgence, Western sweet & fatty, restrained indulgence, and plant-based & alcohol-rich diets. These patterns reflect contemporary, diverse eating behaviours and extend prior research that focused primarily on single nutrients or dietary guideline adherence. Dietary patterns varied by sex, age, language region, education, Swiss-SEP, having children, living situation, and smoking status. Established CVD is relatively uncommon in young participants and we found no evidence of associations between dietary patterns and CVD risk profiles.

Comparing our results with the SJLIFE and general Swiss population data highlights both similarities and unique features of dietary behaviours in Central European/Swiss CCSs. Our findings broadly align with those from the SJLIFE cohort, which identified four predominant dietary patterns among American CCSs, with the largest cluster characterised by fast-food and Western contemporary diets [4]. Identifying five rather than four patterns in our study may indicate greater behavioural heterogeneity within Central European/Swiss CCSs, potentially influenced by diverse dietary habits, although this remains speculative. The number of clusters identified may have been influenced by our sample size, while different clustering approaches and model specifications can yield varying cluster solutions. The similarity between our Western sweet & fatty and Western indulgence patterns and those identified in the Swiss national nutrition survey (menuCH) [20] indicates that Westernised dietary behaviours, characterised by high intakes of meat and processed foods together with both soft or alcoholic drinks, are common among adults in Switzerland. This finding suggests that

CCSs are embedded within the same prevailing dietary environment as the general adult population, rather than adopting distinct survivorship-specific eating patterns.

Multiple sociodemographic and lifestyle determinants were associated with dietary behaviours, which may suggest ways for targeted interventions. Female and higher-educated CCSs more often followed Mediterranean-inspired or plant-based patterns, consistent with evidence linking sex and health literacy to healthier diets [21, 22]. Conversely, men and CCSs with lower educational attainment more frequently adopted Westernised diets. In our study, higher Swiss-SEP was associated not only with a plant-based dietary pattern but also with Western indulgence patterns in CCSs, both included higher alcohol consumption. While higher socioeconomic status is generally associated with higher consumption of fruits, vegetables, and nutrient-rich foods [23], this does not capture alcohol intake. In Switzerland, socioeconomic differences in alcohol use are primarily reflected in drinking patterns and contexts [24], which may partly explain our findings. Consistent with our results, evidence from the Swiss general population shows that lower household income (<6000 CHF/month) is associated with lower adherence to both Western (high intakes of meat and processed foods together with soft drinks or alcohol) and prudent (high intake of fruits, vegetables, white meat and fish) dietary patterns [20]. These findings suggest that socioeconomic differences in dietary patterns do not follow a simple gradient from healthier to less healthy diets. We found that CCSs living with others more likely followed Western indulgence patterns, while having children was associated with reduced alcohol-rich dietary behaviours. This may reflect shared household food environments and caregiving roles, with cohabitation linked to more social eating and childcare responsibilities limiting alcohol consumption [25]. Regional cultural influences were evident, with French- and Italian-speaking CCSs reporting less Westernised diets than those in German-speaking regions [26]. We found that smoking was associated with less healthful dietary patterns, consistent with previous findings [27], highlighting the need for integrated lifestyle interventions addressing multiple risk behaviours simultaneously. Understanding these determinants can help guide targeted interventions aimed at modifying high-risk dietary behaviours among CCSs.

In our study, we did not observe associations between dietary patterns and CVD or CVD risk factors among CCSs. This is broadly consistent with findings from the St. Jude Lifetime Cohort, where greater adherence to healthy dietary patterns, including dietary approaches to stop hypertension (DASH) and Mediterranean diets, was associated with a lower, though not consistently significant, risk of CVD among adult CCSs [28]. A systematic review reported that poorer diets characterised by higher intake of ultra-processed foods or more pro-inflammatory profiles were associated with adverse CVD risk factors, whereas greater adherence to Mediterranean or DASH dietary patterns were associated with more favourable cardiometabolic factors [2]. The absence of associations in our study may partly be explained by the relatively young age of our cohort (median age 34.6 years), with 71% participants younger than 40 years, and the small number of CCSs with CVD. Although CCSs are at increased long-term cardiovascular risk, clinically manifest CVD often develops decades after exposure to cardiotoxic treatments [29]. Therefore, potential effects of dietary patterns on cardiovascular health outcomes may not yet be fully apparent in our participants. Given the progressive nature of CVD risk in CCSs, dietary behaviours established in early adulthood may still influence long-term cardiometabolic trajectories. Longitudinal studies are needed to determine whether modifying dietary patterns can prevent progression from CVD risk factors to CVD and to inform the development of CCSs’ specific dietary guidelines, which are currently lacking.

Our study had several limitations. Our study has a cross-sectional design, which prevents causal inference. CCSs with CVD or CVD risk factors could have adapted their dietary pattern to a more healthful one. Reliance on self-reported diet may underestimate unhealthy and overestimate healthy intake and self-reported CVD and CVD risk factors. These may influence both the distribution of dietary patterns and the observed associations with CVD risk. Last, small sample size reduced our power to detect associations between dietary patterns and CVD risk profiles. Our study was strengthened by its national coverage and the use of a culturally validated FFQ, which identified dietary patterns across the different language regions. We had access to high quality clinical information extracted from the ChCR. The questionnaires gave us access to a wide variety of socio-demographic, and lifestyle factors.

CCSs experience an earlier onset of therapy-related cardiometabolic vulnerability [1], which may amplify the long-term adverse health effects of dietary patterns across the life course. Identifying and modifying adverse dietary patterns, particularly Western indulgence, and understanding the sociodemographic and lifestyle determinants of dietary behaviours in CCSs may be important for informing targeted nutritional interventions. The absence of observable associations in early adulthood should not lead to deprioritising dietary assessment in CCSs follow-up care. Instead, diet may need to be considered as part of a broader, multifactorial risk management strategy rather than as an isolated determinant of cardiovascular risk. Integrating dietary counselling into long-term survivorship care remains important, while tailored dietary recommendations for CCSs still need to be established.

## 5. Conclusion

In conclusion, CCSs exhibit distinct dietary patterns with various nutritional and cardiovascular implications. We did not observe associations between dietary patterns and CVD or CVD risk factors, possibly because of the relatively young age of CCSs, the long latency of clinically manifest disease, and the potential change in diet after the development of CVD or its risk factors. Nevertheless, dietary behaviours remain an important modifiable factor in long-term survivorship care, and dietary counselling should therefore be emphasized. Longitudinal studies are needed to clarify these relationships and to inform the development of tailored dietary recommendations for CCSs.

## Abbreviations

AI: Adequate intake
BMI: Body mass index
CCSs: Childhood cancer survivors
ChCR: Swiss Childhood Cancer Registry
CI: Confidence intervals
CVD: Cardiovascular diseases
DASH: Dietary Approaches to Stop Hypertension
EFSA: European Food Safety Authority
FFQ: Food frequency questionnaire
Gy: Gray
HRQoL: Health-related quality of life
IQR: Interquartile range
N: Number
OR: Odds ratios
PA: Physical activity
PRI: Population reference intake
SCCSS: Swiss Childhood Cancer Survivor Study
SF-36: 36-Item Short Form Survey
Swiss-SEP: Swiss socioeconomic position

## Data Availability

The datasets generated during and/or analysed during the current study are available from the corresponding author on reasonable request.

## Ethics approval

This study received ethical approval from the Ethics Committee of the Canton of Bern, Switzerland (KEK-BE: 166/2014 and 2021-01462).

## Author contributions

**Ruijie Li:** Conceptualization, Formal analysis, Methodology, Software, Validation, Visualization, Writing - original draft, Writing - review & editing. **Raquel Revuelta Iniesta:** Conceptualization, Methodology, Supervision, Writing - review & editing. **Alan R. Barker:** Supervision, Writing - review & editing. **Angeline Chatelan:** Methodology, Writing - review & editing. **Lorenz Leuenberger:** Writing - review & editing. **Yara Shoman:** Writing - review & editing. **Ben Spycher:** Methodology, Writing - review & editing. **Fabiën N. Belle:** Conceptualization, Funding acquisition, Methodology, Project administration, Resources, Supervision, Investigation, Writing - review & editing.

## Acknowledgements

The authors express their gratitude to all childhood cancer survivors for supporting this study. We thank the study team of the Swiss Childhood Cancer Survivor Study, the data managers of the Swiss Paediatric Oncology Group, and the team of the Swiss Childhood Cancer Registry. Finally, we would like to thank Kali Tali for her editorial assistance.

## References

1. Armenian, S.H., et al., Cardiovascular Disease in Survivors of Childhood Cancer: Insights Into Epidemiology, Pathophysiology, and Prevention. J Clin Oncol, 2018. 36(21): p. 2135–2144.

2. Li, R., et al., The Role of Diet in the Cardiovascular Health of Childhood Cancer Survivors-A Systematic Review. Nutrients, 2024. 16(9).

3. Feit, T., et al., Nutritional assessment and dietary intervention among survivors of childhood cancer: current landscape and a look to the future. Front Nutr, 2023. 10: p. 1343104.

4. Lan, T., et al., Dietary patterns and their associations with sociodemographic and lifestyle factors in adult survivors of childhood cancer: a cross-sectional study. Am J Clin Nutr, 2024. 119(3): p. 639–648.

5. Nestel, P.J. and T.A. Mori, Dietary patterns, dietary nutrients and cardiovascular disease. Rev Cardiovasc Med, 2022. 23(1): p. 17.

6. Belle, F.N., et al., Nutritional Assessment of Childhood Cancer Survivors (the Swiss Childhood Cancer Survivor Study-Nutrition): Protocol for a Multicenter Observational Study. JMIR Res Protoc, 2019. 8(11): p. e14427.

7. von Elm, E., Altman, D. G., Egger, M., Pocock, S. J., Gøtzsche, P. C., Vandenbroucke, J. P., & STROBE Initiative., Strengthening the Reporting of Observational Studies in Epidemiology (STROBE) statement: guidelines for reporting observational studies. BMJ (Clinical research ed.), 2007. 335(7624): p. 806–808.

8. Marques-Vidal, P., et al., Dietary intake of subjects with diabetes is inadequate in Switzerland: the CoLaus study. European Journal of Nutrition, 2016. 56(3): p. 981–989.

9. Bernstein, L.H., I.; Morabia, A., Amélioration des performances d’un questionnaire alimentaire semi-quantitatif comparé à un rappel des 24 h (improvement of the effectiveness of a food frequency questionnaire compared with a 24 h recall survey). Santé Publ., 1995(7): p. 403–413.

10. Beer-Borst, S.C., M.; Pechere-Bertschi, A.; Morabia, A., Twelve-year trends and correlates of dietary salt intakes for the general adult population of geneva, switzerland. Eur. J. Clin. Nutr., 2009(63): p. 155.

11. Morabia, A.B., M.; Kumanyika, S.; Sorenson, A.; Mabiala, I.; Prodolliet, B.; Rolfo, I.; Luong, B., Development and validation of a semi-quantitative food questionnaire based on a population survey. Sozial Praventivmedizin 1994(39): p. 345–369

12. World Health Organization. Obesity: preventing and managing the global epidemic. Report of a WHO consultation. Tech Rep Ser. 2000;894:i–xii, 1-253.

13. Bull, F.C., et al., World Health Organization 2020 guidelines on physical activity and sedentary behaviour. Br J Sports Med, 2020. 54(24): p. 1451–1462.

14. Panczak, R., et al., The Swiss neighbourhood index of socioeconomic position: update and re-validation. Swiss Med Wkly, 2023. 153: p. 40028.

15. RAND Health Care. 36-Item Short Form Survey (SF-36) Scoring Instructions. 2025. Available from: https://www.rand.org/health-care/surveys_tools/mos/36-item-short-form/scoring.html.

16. Steliarova-Foucher, E., et al., International Classification of Childhood Cancer, third edition. Cancer, 2005. 103(7): p. 1457–67.

17. Children’s Oncology Group (COG). Long-Term Follow-Up Guidelines for Survivors of Childhood, Adolescent and Young Adult Cancers, Version 6.0. 2023.

18. Belle, F.N., et al., No evidence of overweight in long-term survivors of childhood cancer after glucocorticoid treatment. Cancer, 2018. 124(17): p. 3576–3585.

19. Office fédéral de la sécurité alimentaire et des affaires vétérinaires. Des valeurs nutritionnelles de référence suisses. 2024. Available from: https://www.blv.admin.ch/blv/fr/home/lebensmittel-und-ernaehrung/ernaehrung/empfehlungen-informationen/naehrstoffe/naehrstoffzufuhr-dynamische-tabelle.html.

20. Krieger, J.P., et al., Dietary Patterns and Their Sociodemographic and Lifestyle Determinants in Switzerland: Results from the National Nutrition Survey menuCH. Nutrients, 2018. 11(1).

21. Wardle, J., Haase, A. M., Steptoe, A., Nillapun, M., Jonwutiwes, K., & Bellisle, F., Gender differences in food choice: the contribution of health beliefs and dieting. Ann Behav Med, 2004. 27 (2): p. 107–116

22. Klink, U., et al., Socioeconomic differences in animal food consumption: Education rather than income makes a difference. Front Nutr, 2022. 9: p. 993379.

23. Darmon, N. and A. Drewnowski, Does social class predict diet quality? Am J Clin Nutr, 2008. 87(5): p. 1107–17.

24. Sandoval, J.L., et al., Alcohol control policies and socioeconomic inequalities in hazardous alcohol consumption: a 22-year cross-sectional study in a Swiss urban population. BMJ Open, 2019. 9(5): p. e028971.

25. Pickard, A., et al., Associations between parent and child latent eating profiles and the role of parental feeding practices. Appetite, 2024. 201: p. 107589.

26. Chatelan, A., et al., Major Differences in Diet across Three Linguistic Regions of Switzerland: Results from the First National Nutrition Survey menuCH. Nutrients, 2017. 9(11).

27. Spring, B., A.C. Moller, and M.J. Coons, Multiple health behaviours: overview and implications. J Public Health (Oxf), 2012. 34 **Suppl 1**(Suppl 1): p. i3–10.

28. Lan, T., et al., Adherence to healthy diet and risk of cardiovascular disease in adult survivors of childhood cancer in the St. Jude Lifetime Cohort: a cross-sectional study. BMC Med, 2023. 21(1): p. 242.

29. Mulrooney, D.A., et al., Cardiac outcomes in a cohort of adult survivors of childhood and adolescent cancer: retrospective analysis of the Childhood Cancer Survivor Study cohort. BMJ, 2009. 339: p. b4606.

