## Supplementary Material for "Dietary Patterns and Cardiovascular Risk in Adult Survivors of Childhood Cancer: Findings From the SCCSS-Nutrition Study"

Supplementary Figure 1. Response rates in the Swiss Childhood Cancer Survivor Study (SCCSS)-Nutrition study.

Supplementary Table 1. Food group items in adult childhood cancer survivors: Swiss Childhood Cancer Survivor Study (SCCSS)- Nutrition study.

Supplementary Table 2. Comparison of sociodemographic and clinical characteristics among included and excluded participants in the Swiss Childhood Cancer Survivor Study (SCCSS)- Nutrition study.

Supplementary Table 3. Median consumption for each food group by diet patterns in adult childhood cancer survivors in Switzerland.

Supplementary Table 4. Median macro- and micronutrient intake by dietary patterns in adult childhood cancer survivors.

Supplementary Table 5. Socio-demographic, lifestyle, and clinical characteristics of adult childhood cancer survivors across the five dietary patterns (univariable multinomial logistic regressions).

Supplementary Table 6. Association of sociodemographic, lifestyle and clinical characteristics by CVD risk profiles (univariable multinomial logistic regressions).

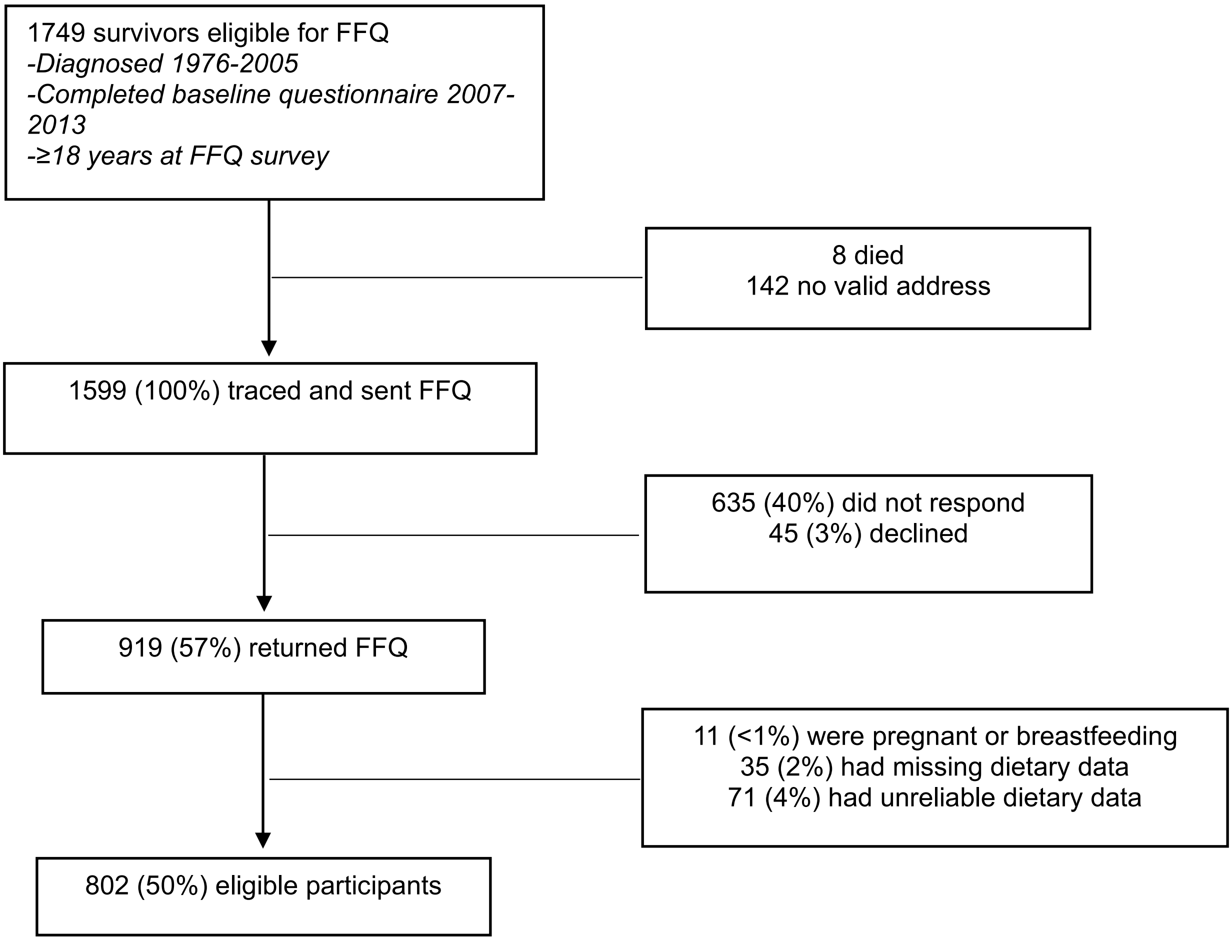


Supplementary Figure 1. Response rates in the Swiss Childhood Cancer Survivor Study (SCCSS)-Nutrition study.

*FFQ, food frequency questionnaire*

Supplementary Table 1. Food group items in adult childhood cancer survivors: Swiss Childhood Cancer Survivor Study (SCCSS)- Nutrition study.

| Foods or food groups | Food items |
|---|---|
| Whole grains | Wholemeal bread, rye bread, muesli or other mixed cereals |
| Refined and non-whole grains | White bread, sliced bread, farmhouse bread, milk bread, plaited bread, cornflakes, puffed wheat, puffed cereals, rusks, crackers, Swedish toast |
| Rice and couscous | Rice, wheat semolina, couscous |
| Fresh fruits | Banana, apple, pear, plum, grape, orange, tangerine, clementine, peach, nectarine, apricot, melon, strawberries, raspberries, blueberries, blackcurrants, kiwi fruit |
| Preserved fruits | Tinned fruit, sweet compote |
| Green leafy vegetables | Green beans, spinach, cauliflower, broccoli, green salad |
| Fruiting vegetables | Tomatoes, avocado, cucumbers, zucchini, eggplant, bell peppers |
| Root vegetables | Carrots |
| Other vegetables | Peas, corn, onions, mushrooms |
| Potato | Plain potatoes |
| French fries | French fries |
| Clear vegetable soup | Clear vegetable soup (vegetable stock) |
| Thick vegetable soup | Thick vegetable soup (with peas, beans, minestrone) |
| Tofu | Tofu |
| Nuts | Nuts (almonds, walnuts, peanuts, hazelnuts, etc.) |
| Low-fat dairy | Plain yoghurt, light yoghurt, light custard, light cream, 0% fromage frais, curd cheese 20%, ricotta, sere, cottage cheese, skimmed milk (0%) in coffee, skimmed (0%) milk for drinking |
| High-fat dairy | Fruit yoghurt, custard dessert, cream, Feta, mozzarella, fromage frais 1/2 salt, petit-suisse, Gruyere, tomme, camembert, blue, parmesan, cheese fondue, 35% whole cream, whole or semi-skimmed milk in coffee, coffee creamer, whole or semi-skimmed milk for drinking, other cheeses (Emmental, raclette, etc.), Swiss-style muesli with oats, grated apple, nuts, and yogurt (Birchermüesli) |
| Processed meat | Sausage, cured ham, bacon, salami, pasta terrine, saveloy |
| Red meat | Beefsteak, horse, veal (escalope, fillet), chopped steak, entrecôte, röti (beef, pork, veal), ham, pot-au-feu, lamb chops, pork chops |
| Organ meats | Liver (veal, beef, and pork) |
| Poultry | Chicken without skin, chicken with skin |
| White fish | Lean fish such as cod, hake, trout |
| Fried fish | Fried or breaded fish (perch fillets) |
| Fatty fish | Fresh salmon, smoked salmon, tuna in oil |
| Shellfish | Prawns, other shellfish |
| Pastries | Croissant, chocolate croissant |
| Sweets and dessert | Fruit tart, cream cake, cake, dry pastry, biscuits, cookies, chocolate, ice cream, sorbet |
| Sugar and sweeteners | Artificial sugar (assugrin, aspartam), sugar |
| Jam, honey and Nutella | Jam, honey, and Nutella |
| Pasta | Pasta products, ravioli, tortellini, cannelloni |
| Pizza | Pizza |
| Quiche Lorraine | Quiche Lorraine |
| Eggs | Eggs |
| Salty snacks | Crisps |
| Olives | Black and green olives |
| Water | Aproz, Valser, San Pellegrino, Passuger, Perrier, Vitel, Volvic, tap water, Henniez, |

| | |
|---|---|
| | Evian, Vichy |
| Coffee | Coffee |
| Tea | Tea, infusion |
| Sweetened beverages | Lemonade, cola, soda, fruit syrup, fruit juice in bottles or cartons |
| Protein shakes | Hot or cold chocolate, milk-based mixed drinks, protein shakes |
| Fresh fruit juices | Fresh fruit juices |
| Beer | Beer |
| Wine and champagne | Wine and champagne |
| Spirits | Anisette and martini aperitifs, strong alcohol such as whisky, brandy, or liqueur |
| Margarine | Low-fat margarine, Margarine |
| Butter | Butter |
| Oil | Olive oil, peanut oil, sunflower oil, other oils e.g., rapeseed oil |
| Vitamin and mineral supplements | Vitamin C, Vitamin E, Multivitamins, Son, Garlic tablets |
| Sauces | Mayonnaise, fatty sauces e.g., cream sauce, sauce with coconut milk |
| Bolognese and tomato sauce | Bolognese sauce, tomato sauce |
| Salt | Added salt |
| Vinegar | Vinegar |

Supplementary Table 2. Comparison of sociodemographic and clinical characteristics among included and excluded participants in the Swiss Childhood Cancer Survivor Study (SCCSS)- Nutrition study.

| | Included participants (N = 802) | Excluded participants (N = 117) | *p* - value[a] |
|---|---|---|---|
| Sex | | | |
| Female | 401 (50) | 57 (49) | 0.796 |
| Male | 401 (50) | 60 (51) | |
| Age at survey, years | | | |
| Median (IQR) | 34.6 (28.8;41.1) | 32.8 (28.1;39.6) | 0.138 |
| ≤30 | 248 (31) | 44 (38) | 0.249 |
| 31-39 | 320 (40) | 46 (39) | |
| ≥40 | 234 (29) | 27 (23) | |
| Language region within Switzerland | | | |
| German speaking | 570 (71) | 84 (71) | 0.931 |
| French speaking | 214 (27) | 31 (27) | |
| Italian speaking | 18 (2) | 2 (2) | |
| Migration background | | | |
| Swiss, no migration background | 635 (79) | 91 (78) | 0.654 |
| Migration background | 163 (20) | 26 (22) | |
| Missing | 4 (<1) | - | |
| Education (highest degree) | | | |
| Lower than university | 532 (66) | 86 (74) | 0.123 |
| University | 270 (34) | 31 (27) | |
| Swiss-SEP[c] | | | |
| Median (IQR) | 62.1 (55.3;69.2) | 63.6 (57.9;72.1) | 0.061 |
| Low | 306 (38) | 32 (27) | 0.075 |
| Medium | 272 (34) | 41 (35) | |
| High | 217 (27) | 40 (34) | |
| Missing | 7 (<1) | 4 (3) | |
| Marital status | | | |
| Married | 287 (36) | 36 (31) | 0.291 |
| Single & Widowed & Divorced | 509 (63) | 80 (68) | |
| Missing | 6 (<1) | 1 (<1) | |
| Children | | | |
| No | 533 (66) | 88 (75) | 0.059 |
| Yes | 269 (34) | 29 (25) | |
| Living situation | | | |
| Alone | 164 (20) | 23 (20) | 0.946 |
| With others | 638 (80) | 91 (78) | |
| Missing | - | 3 (3) | |

| | | | |
|---|---|---|---|
| Smoking status | | | |
| Never | 542 (68) | 74 (67) | 0.921 |
| Former | 132 (16) | 17 (15) | |
| Current | 128 (16) | 19 (17) | |
| Physical activity[c] | | | |
| Inactive | 171 (21) | 42 (36) | **<0.001** |
| Active | 629 (78) | 74 (63) | |
| Missing | 2 (<1) | 1 (<1) | |
| SF-36[d], mean ± SD | | | |
| Physical functioning | 51.07 ± 8.97 | 50.22 ± 10.67 | 0.387 |
| Role physical | 51.74 ± 9.48 | 50.06 ± 11.75 | 0.346 |
| Bodily pain | 55.24 ± 6.65 | 54.81 ± 8.18 | 0.719 |
| General health | 50.90 ± 11.87 | 48.96 ± 12.87 | 0.178 |
| Vitality | 49.40 ± 10.36 | 48.64 ± 12.36 | 0.826 |
| Social functioning | 50.49 ± 10.40 | 49.53 ± 11.20 | 0.342 |
| Role emotional | 50.33 ± 10.19 | 48.89 ± 12.55 | 0.464 |
| Mental health | 40.59 ± 8.05 | 40.43 ± 8.81 | 0.893 |
| Physical component summary | 55.19 ± 8.68 | 54.03 ± 11.04 | 0.512 |
| Mental component summary | 45.31 ± 9.45 | 44.43 ± 10.53 | 0.669 |
| BMI at survey | | | |
| Underweight, <18.5 kg/m$^2$ | 39 (5) | 6 (5) | **0.040** |
| Normal, 18.5 to <25 kg/m$^2$ | 511 (64) | 61 (52) | |
| Overweight, ≥25 to <30 kg/m$^2$ | 177 (22) | 24 (21) | |
| Obese, ≥30 kg/m$^2$ | 75 (9) | 20 (17) | |
| Missing | - | 6 (5) | |
| ICCC-3 cancer diagnosis | | | |
| I Leukaemias | 246 (31) | 33 (28) | **0.046** |
| II Lymphomas | 173 (22) | 17 (15) | |
| III Central nervous system tumours | 81 (10) | 23 (20) | |
| IV Neuroblastoma | 28 (3) | 3 (3) | |
| V Retinoblastoma | 12 (2) | 4 (3) | |
| VI Renal tumours | 52 (6) | 6 (5) | |
| VII Hepatic tumours | 6 (<1) | 1 (<1) | |
| VIII Malignant bone tumours | 50 (6) | 3 (3) | |
| IX Soft tissue sarcomas | 66 (8) | 9 (8) | |
| X Germ cell tumours | 43 (5) | 6 (5) | |
| XI & XII Other tumours | 26 (3) | 6 (5) | |
| Langerhans Cell Histiocytosis | 19 (2) | 6 (5) | |
| Age at diagnosis, years | | | |
| Median (IQR) | 9.7 (3.9;13.9) | 6.4 (3.3;13.3) | 0.061 |

| | | | |
|---|---|---|---|
| <5 | 251 (31) | 44 (38) | 0.115 |
| 5-9 | 164 (20) | 30 (26) | |
| 10-14 | 239 (30) | 24 (21) | |
| ≥15 | 148 (18) | 19 (16) | |
| Time since diagnosis, years | | | |
| Median (IQR) | 26.1 (20.2;31.7) | 25.2 (20.1;31.3) | 0.845 |
| ≤25 | 377 (47) | 62 (53) | 0.226 |
| ≥25 | 425 (53) | 55 (47) | |
| History of relapse, yes | 99 (12) | 16 (14) | 0.684 |
| HSCT, yes | 30 (4) | 8 (7) | 0.116 |
| Cranial radiation, yes | 131 (16) | 24 (21) | 0.259 |
| <18Gy | 38 (5) | 10 (9) | 0.216 |
| ≥18Gy | 93 (12) | 14 (12) | |
| Chest radiation, yes | 93 (12) | 16 (14) | 0.516 |
| <30 Gy | 36 (4) | 7 (6) | 0.703 |
| ≥30 Gy | 57 (7) | 9 (8) | |
| TBI and/or abdominal radiation, yes | 71 (9) | 14 (12) | 0.278 |
| Anthracyclines, yes | 298 (37) | 38 (32) | 0.326 |
| Glucocorticoids, yes | 339 (42) | 50 (43) | 0.725 |
| Alkylating agents, yes | 331 (41) | 47 (40) | 0.821 |

*Abbreviations: BMI, body mass index; Gy, gray; HSCT, hematopoietic stem cell transplantation; ICCC-3, International Classification of Childhood Cancer 3rd edition; IQR, interquartile range; N, number; SD, standard deviation; SEP, socioeconomic position; SF-36, 36-Item Short Form Survey; TBI, total body irradiation. Data are presented as N (%) or median (IQR).*

[a] *p* - value was calculated using chi-square test for categorical variables or the Kruskal-Wallis test for continuous variables.

[b] Swiss neighbourhood index of socioeconomic position (Swiss-SEP) is a validated area-based index (range 0–100) reflecting household education and occupation, housing conditions, and rent values. Values were categorised into tertiles (low, medium, high) based on the national Swiss population distribution.

[c] PA active: ≥150 min of moderate intense or 75 min of vigorous intense or a combination of moderate and vigorous intense physical activity per week.

[d] SF-36, all scores are norm-based (mean = 50, SD = 10). Higher scores indicate better health-related quality of life. The physical component summary score is primarily derived from physical functioning, role physical, bodily pain, and general health domains, whereas the mental component summary score is primarily derived from vitality, social functioning, role emotional, and mental health domains.

Supplementary Table 3. Median consumption for each food group by diet patterns in adult childhood cancer survivors in Switzerland.

| Daily intake (g/1000kcal) Median (IQR) | Total (N = 802) | Diet patterns | | | | |
|---|---|---|---|---|---|---|
| | | Mediterranean-Inspired Diet (N = 174) | Western Indulgence Diet (N = 304) | Western Sweet & Fatty Diet (N = 207) | Restrained Indulgence Diet (N = 71) | Plant-based & Alcohol-rich Diet (N = 46) |
| Whole grains | 13.75 (4.48;29.85) | 20.69 (7.82;35.39) | 11.39 (3.2;23.88) | 16.81 (4.92;30.87) | 8.16 (0;41.68) | 19.06 (10.55;38.69) |
| Refined and non-whole grains | 14.57 (6.85;31.31) | 16.22 (9.02;38.93) | 13.22 (7.10;26.94) | 18.48 (9.14;38.14) | 5.83 (0;21.61) | 11.98 (5.34;29.21) |
| Rice and couscous | 17.36 (9.36;26.12) | 21.82 (10.94;32.58) | 16.68 (9.45;24.43) | 15.65 (9.27;23.61) | 6.72 (0;21.77) | 26.06 (15.62;36.52) |
| Fresh fruits | 73.36 (30.93;152.99) | 126.34 (62.37;206.61) | 57.98 (26.33;115.63) | 61.85 (27.06;113.96) | 70.70 (7.59;171.13) | 121.71 (55.99;246.49) |
| Preserved fruits | 0 (0;0) | 0 (0;0) | 0 (0;0) | 0 (0;1.32) | 0 (0;0) | 0 (0;1.27) |
| Green leafy vegetables | 58.37 (34.60;91.23) | 60.34 (39.81;95.10) | 55.26 (31.59;85.16) | 56.29 (34.60;88.06) | 59.92 (15.87;109.53) | 90.83 (64.63;125.76) |
| Fruiting vegetables | 49.95 (24.30;82.70) | 55.35 (29.72;89.30) | 50.53 (28.99;82.84) | 33.23 (13.70;62.64) | 56.20 (25.51;76.91) | 97.08 (26.57;126.96) |
| Root vegetables | 13.49 (6.76;24.37) | 16.21 (8.37;30.08) | 11.80 (6.23;19.76) | 12.68 (6.15;25.78) | 8.81 (2.19;22.93) | 26.48 (15.88;53.21) |
| Other vegetables | 34.54 (18.45;56.91) | 31.99 (15.82;57.82) | 37.72 (22.28;58.39) | 30.14 (12.87;50.10) | 32.01 (14.42;53.42) | 52.94 (38.36;75.18) |
| Potato | 15.92 (8.21;26.25) | 13.91 (7.52;25.56) | 16.22 (8.63;25.67) | 17.77 (8.63;29.26) | 8.86 (0;17.27) | 18.44 (12.36;31.06) |
| French fries | 3.62 (1.42;7.65) | 2.60 (0;5.42) | 5.31 (2.63;9.13) | 3.75 (2.06;8.01) | 0 (0;3.87) | 2.68 (0;4.83) |
| Clear vegetable soup | 5.87 (0;16.40) | 6.42 (0;20.30) | 5.72 (0;14.06) | 4.96 (0;13.43) | 0 (0;19.47) | 18.45 (4.85;43.00) |
| Thick vegetable soup | 6.79 (0;18.60) | 7.06 (0;20.49) | 6.07 (0;16.63) | 7.95 (0;19.68) | 0 (0;10.68) | 21.31 (0;55.13) |
| Tofu | 0 (0;0) | 0 (0;0) | 0 (0;0) | 0 (0;0) | 0 (0;0) | 6.67 (0;11.89) |
| Nuts | 1.29 (0.31;3.71) | 1.32 (0;4.32) | 1.24 (0.43;3.14) | 0.28 (1.12;2.99) | 1.35 (0;9.25) | 2.35 (0.79;5.49) |
| Low-fat dairy | 11.19 (1.84;37.82) | 19.68 (3.73;61.90) | 8.21 (0;28.73) | 9.64 (3.05;30.07) | 11.73 (0;103.99) | 9.13 (0;44.91) |
| High-fat dairy | 74.75 (40.76;136.50) | 59.48 (35.02;114.94) | 60.44 (37.48;103.31) | 118.56 (61.57;186.27) | 95.20 (41.32;188.23) | 71.53 (34.60;116.34) |
| Processed meat | 8.20 (3.19;15.73) | 5.71 (1.93;12.36) | 10.45 (4.90;18.15) | 9.01 (3.91;17.41) | 7.05 (1.26;19.27) | 0 (0;4.25) |
| Red meat | 22.11 (11.15;35.75) | 16.54 (7.70;30.89) | 28.29 (17.79;42.83) | 20.43 (11.47;33.33) | 23.03 (12.32;35.82) | 0.86 (0;17.14) |
| Organ meats | 0 (0;0) | 0 (0;0) | 0 (0;0) | 0 (0;0) | 0 (0;0) | 0 (0;0) |
| Poultry | 9.08 (4.35;15.68) | 8.93 (4.33;15.93) | 10.50 (5.75;17.11) | 7.33 (3.90;12.82) | 13.01 (5.47;19.10) | 0 (0;4.03) |
| White fish | 0 (0;5.52) | 3.66 (0;9.72) | 2.37 (0;5.65) | 0 (0;2.74) | 0 (0;8.86) | 0 (0;2.96) |
| Fried fish | 0.60 (0;4.52) | 2.51 (0;5.28) | 2.53 (0;4.74) | 1.75 (0;4.52) | 0 (0;3.66) | 0 (0;0) |

| | | | | | | |
|---|---|---|---|---|---|---|
| Fatty fish | 2.90 (0.41;5.96) | 3.77 (1.49;8.20) | 3.69 (1.74;6.41) | 1.67 (0;3.51) | 3.32 (0;7.25) | 0 (0;2.74) |
| Shellfish | 0 (0;2.08) | 1.50 (0;3.65) | 0 (0;2.39) | 0 (0;0) | 0 (0;2.66) | 0 (0;0) |
| Pastries | 1.38 (0;3.14) | 1.22 (0;3.2) | 1.75 (0.77;4.23) | 0 (1.32;2.63) | 0 (0;0.74) | 1.82 (0;3.76) |
| Sweets and dessert | 15.90 (8.62;25.66) | 17.52 (10.51;27.02) | 14.94 (7.91;23.78) | 17.66 (10.30;29.55) | 7.91 (1.25;20.27) | 16.04 (9.33;21.98) |
| Sugar and sweeteners | 0.51 (0;3.42) | 0.12 (0;1.72) | 0.53 (0;2.53) | 2.21 (0;4.98) | 0 (0;0.70) | 0.97 (0;3.88) |
| Jam, honey and Nutella | 3.15 (1.05;7.20) | 4.84 (1.77;9.48) | 2.17 (0.60;4.52) | 5.77 (2.41;9.66) | 0.83 (0;2.81) | 2.96 (0.92;5.19) |
| Pasta | 36.32 (21.13;55.86) | 32.51 (21.27;52.88) | 39.62 (24.23;57.87) | 35.58 (22.34;56.07) | 12.96 (0;36.88) | 49.08 (22.81;59.83) |
| Pizza | 10.52 (5.80;18.38) | 8.60 (5.07;15.58) | 13.95 (7.62;21.69) | 9.75 (5.96;17.56) | 4.00 (0;14.75) | 8.62 (5.70;17.23) |
| Quiche Lorraine | 0 (0;1.77) | 0 (0;0) | 0 (0;2.11) | 0 (0;1.62) | 0 (0;0) | 0 (0;3.50) |
| Eggs | 9.93 (4.90;18.16) | 7.63 (3.59;13.98) | 11.26 (6.03;19.58) | 10.42 (5.58;16.99) | 13.37 (0;24.79) | 13.37 (2.21;19.62) |
| Salty snacks | 0.80 (0;1.69) | 0.51 (0;0.94) | 1.20 (0.57;2.14) | 0.88 (0.44;1.77) | 0.54 (0;1.41) | 0.73 (0;1.82) |
| Olives | 0.25 (0;0.75) | 0.28 (0;0.78) | 0.42 (0;0.96) | 0 (0;0.41) | 0 (0;0.67) | 0.37 (0;0.92) |
| Water | 455.72 (285.67;655.28) | 458.15 (323.43;654.84) | 477.60 (307.47;676.48) | 449.67 (270.45;640.80) | 379.86 (10.95;676.79) | 443.11 (271.86;616.86) |
| Coffee | 66.46 (10.72;115.18) | 58.79 (5.04;106.46) | 85.08 (38.15;129.78) | 51.82 (4.40;95.53) | 23.78 (0;111.95) | 81.39 (26.75;120.10) |
| Tea | 23.81 (0;115.10) | 75.15 (6.91;195.68) | 11.34 (0;51.42) | 36.43 (3.74;128.86) | 0 (0;50.56) | 111.58 (23.85;211.06) |
| Sweetened beverages | 36.98 (9.58;115.51) | 21.59 (3.41;83.12) | 35.93 (13.24;102.01) | 70.20 (21.66;144.87) | 11.34 (0;118.26) | 23.04 (5.15;83.96) |
| Protein shakes | 0 (0;18.64) | 0 (0;6.82) | 0 (0;15.20) | 8.93 (0;34.10) | 3.96 (0;42.74) | 0 (0;11.82) |
| Fresh fruit juices | 4.58 (0;20.00) | 8.60 (0;35.72) | 6.79 (0;21.10) | 2.45 (0;12.91) | 0 (0;4.64) | 7.71 (0;19.74) |
| Beer | 7.53 (0;36.17) | 0 (0;10.68) | 32.15 (7.37;58.84) | 0 (0;16.07) | 0 (0;11.25) | 17.91 (0;48.50) |
| Wine and champagne | 7.29 (0;19.07) | 3.43 (0;10.52) | 14.01 (6.58;26.91) | 2.89 (0;9.94) | 0 (0;10.46) | 12.02 (5.58;24.78) |
| Spirits | 0 (0;3.55) | 0 (0;1.09) | 3.39 (0;6.10) | 0 (0;1.84) | 0 (0;0) | 1.55 (0;3.50) |
| Margarine | 0 (0;0.14) | 0 (0;2.06) | 0 (0;0.15) | 0 (0;0) | 0 (0;0) | 0 (0;0) |
| Butter | 2.44 (0.85;5.41) | 1.46 (0.50;3.50) | 2.25 (0.93;4.75) | 4.87 (2.41;8.02) | 1.41 (0;4.97) | 1.95 (0.16;4.85) |
| Oil | 3.73 (1.92;6.25) | 4.08 (2.09;6.80) | 3.70 (2.00;5.64) | 3.55 (1.64;5.41) | 3.20 (0.55;6.99) | 5.51 (2.89;8.17) |
| Vitamin and mineral supplements | 0 (0;0.02) | 0 (0;0.04) | 0 (0.05) | 0 (0;0.02) | 0 (0;0.01) | 0 (0;0) |
| Sauces | 2.32 (0.84;4.78) | 1.25 (0.17;2.74) | 3.07 (1.49;5.61) | 2.80 (1.30;5.10) | 1.02 (0;2.67) | 1.90 (0.25;5.01) |
| Bolognese and tomato sauce | 10.25 (5.21;16.60) | 9.05 (4.83;15.13) | 11.90 (6.70;18.05) | 9.31 (5.20;15.74) | 6.08 (1.59;16.11) | 11.10 (4.73;15.29) |
| Salt | 0.06 (0;0.18) | 0.04 (0;0.13) | 0.07 (0.01;0.19) | 0.06 (0;0.19) | 0.02 (0;0.12) | 0.16 (0.05;0.30) |

| | | | | | | |
|---|---|---|---|---|---|---|
| Vinegar | 4.31 (1.58;8.07) | 3.33 (0.86;6.84) | 4.11 (1.78;7.95) | 4.98 (2.15;8.40) | 2.69 (0;9.60) | 7.27 (4.54;10.86) |

*Abbreviations: IQR, interquartile range; N, number.*

Supplementary Table 4. Median macro- and micronutrient intake by dietary patterns in adult childhood cancer survivors.

| Daily intake Median (IQR) | Total (N = 802) | Diet patterns: Mediterranean-Inspired Diet (N = 174) | Western Indulgence Diet (N = 304) | Western Sweet & Fatty Diet (N = 207) | Restrained Indulgence Diet (N = 71) | Plant-based & Alcohol-rich Diet (N = 46) | *p* – value[a] |
|---|---|---|---|---|---|---|---|
| Macronutrients | | | | | | | |
| Total energy, Kcal | 1533.09 (1231.72;1901.84) | 1497.25 (1208.74;1871.90) | 1550.10 (1244.66;1902.87) | 1256.93 (1652.778;2004.19) | 1429.26 (1095.61;1805.05) | 1405.06 (1267.13;1813.73) | **0.023** |
| Total protein, % of energy | 15.88 (13.71;18.04) | 15.91 (13.84;17.72) | 16.44 (14.46;18.76) | 14.97 (13.11;17.20) | 17.21 (13.95;20.66) | 12.90 (10.16;16.04) | **<0.001** |
| Vegetable protein | 4.33 (3.70;5.09) | 4.83 (4.22;5.64) | 4.06 (3.57;4.64) | 4.30 (3.76;4.97) | 4.05 (3.16;4.70) | 5.73 (5.36;6.53) | **<0.001** |
| Animal protein | 11.34 (8.97;13.87) | 10.92 (8.79;12.95) | 12.36 (10.08;15.01) | 10.63 (8.47;12.89) | 13.18 (9.54;16.65) | 6.91 (4.37;9.97) | **<0.001** |
| Carbohydrate, % of energy | 44.15 (38.09;50.21) | 48.36 (43.49;52.84) | 39.78 (35.67;45.20) | 46.48 (40.98;50.98) | 40.22 (34.22;50.89) | 49.19 (42.44;54.63) | **<0.001** |
| Fibre, g | 10.73 (7.76;14.83) | 12.51 (8.95;17.86) | 9.68 (7.15;13.00) | 11.02 (8.44;14.51) | 9.29 (5.58;15.29) | 13.58 (10.47;18.96) | **<0.001** |
| Total fat, % of energy | 37.17 (32.75;41.40) | 34.22 (31.10;39.13) | 38.56 (34.68;42.52) | 37.25 (33.04;41.27) | 39.59 (32.67;45.54) | 35.18 (30.81;39.80) | **<0.001** |
| Saturated fat | 13.62 (11.67;15.82) | 12.50 (10.59;14.07) | 13.85 (12.08;15.85) | 14.42 (12.71;16.55) | 13.64 (11.21;17.24) | 11.89 (9.66;13.92) | **<0.001** |
| Monounsaturated fat | 15.31 (12.97;17.59) | 14.17 (11.90;16.81) | 15.98 (13.98;18.25) | 14.97 (12.64;16.78) | 16.22 (13.14;20.55) | 15.44 (12.58;19.39) | **<0.001** |
| Polyunsaturated fat | 4.60 (3.92;5.43) | 4.63 (3.94;5.52) | 4.82 (4.15;5.48) | 4.20 (3.63;4.79) | 4.81 (3.83;5.80) | 5.21 (3.68;6.24) | **<0.001** |
| Cholesterol, mg | 295.35 (220.83;414.90) | 258.12 (188.70;346.48) | 311.06 (237.47;427.86) | 320.75 (232.00;443.33) | 264.48 (201.28;433.71) | 235.00 (144.90;320.60) | **<0.001** |
| Total sugars, g | 75.02 (52.83;106.96) | 77.88 (57.21;117.50) | 66.92 (47.46;95.69) | 82.58 (63.12;120.10) | 61.43 (44.79;109.33) | 72.09 (57.18;106.66) | **<0.001** |
| Micronutrients | | | | | | | |
| Vitamin A, mg-RE | 0.57 (0.41;0.84) | 0.57 (0.41;0.86) | 0.56 (0.40;0.81) | 0.60 (0.42;0.82) | 0.53 (0.33;0.78) | 0.73 (0.50;0.93) | 0.055 |
| Vitamin D, µg | 1.92 (1.20;2.94) | 2.24 (1.35;3.46) | 2.07 (1.37;3.17) | 1.57 (0.89;2.55) | 2.16 (1.13;3.15) | 1.23 (0.78;1.99) | **<0.001** |
| Sodium, g | 3.22 (2.23;3.50) | 2.33 (2.19;3.44) | 3.43 (2.33;3.53) | 2.59 (2.21;3.50) | 3.30 (2.18;3.46) | 2.34 (2.23;3.14) | **<0.001** |
| Potassium, g | 2.59 (2.00;3.27) | 2.59 (2.06;3.31) | 2.58 (2.00;3.08) | 2.52 (1.99;3.36) | 2.44 (1.75;3.33) | 3.00 (2.15;3.64) | 0.304 |
| Calcium, mg | 784.81 (577.04;1078.44) | 781.00 (544.60;1050.03) | 754.95 (578.54;1020.04) | 818.76 (592.76;1196.19) | 831.06 (534.219;1206.63) | 681.10 (590.94;1063.85) | 0.067 |
| Phosphorus, mg | 1161.65 (910.64;1454.58) | 1146.45 (861.65;1378.77) | 1137.89 (931.73;1434.75) | 1217.41 (898.95;1575.55) | 1123.81 (935.67;1445.56) | 1195.78 (899.61;1464.46) | 0.242 |

| | | | | | | | |
|---|---|---|---|---|---|---|---|
| Iron, mg | 8.36 (6.67;10.59) | 8.10 (6.65;10.67) | 8.82 (6.86;109.94) | 8.32 (6.52;10.35) | 7.85 (5.91;9.79) | 8.22 (6.99;10.26) | 0.098 |
| Others | | | | | | | |
| Total alcohol, g | 3.36 (0.48;8.04) | 1.18 (0;3.51) | 7.43 (4.24;11.43) | 1.68 (0;4.70) | 1.29 (0;3.30) | 4.60 (2.49;9.58) | **<0.001** |

*Abbreviations: IQR, interquartile range; N, number.*

[a] *p* - value was calculated using Kruskal-Wallis test for continuous variables.

Supplementary Table 5. Socio-demographic, lifestyle, and clinical characteristics of adult childhood cancer survivors across the five dietary patterns (univariable multinomial logistic regressions).

| | Compared with a Mediterranean-Inspired Diet (N = 174) | | | | | | | |
|---|---|---|---|---|---|---|---|---|
| | Western Indulgence Diet (N = 304) | | Western Sweet & Fatty Diet (N = 207) | | Restrained Indulgence Diet (N = 71) | | Plant-based & Alcohol-rich Diet (N = 46) | |
| | OR (95% CI) | *p* – value[a] | OR (95% CI) | *p* – value[a] | OR (95% CI) | *p* – value[a] | OR (95% CI) | *p* – value[a] |
| Sex | | | | | | | | |
| Female | (ref) | **<0.001** | (ref) | **0.026** | (ref) | **0.024** | (ref) | 0.155 |
| Male | 3.41 (2.31-5.04) | | 1.60 (1.06-2.42) | | 1.91 (1.09-3.34) | | 0.58 (0.28-1.23) | |
| Age at survey, years | 0.97 (0.95-0.99) | **0.006** | 0.97 (0.94,0.99) | **0.008** | 1.00 (0.96-1.03) | 0.823 | 0.91 (0.87-0.95) | **<0.001** |
| Language region within Switzerland | | | | | | | | |
| German speaking | (ref) | **0.003** | (ref) | **<0.001** | (ref) | 0.121 | (ref) | 0.538 |
| French speaking | 0.56 (0.38-0.85) | | 0.56 (0.38-0.85) | | 0.74 (0.36-1.49) | | 0.44 (0.28-0.69) | |
| Italian speaking | 0.23 (0.07-0.78) | | 0.23 (0.07-0.78) | | 0.42 (0.05-3.45) | | 0.24 (0.06-0.93) | |
| Migration background | | | | | | | | |
| Swiss, no migration background | (ref) | 0.664 | (ref) | 0.327 | (ref) | 0.477 | (ref) | 0.922 |
| Migration background | 0.90 (0.58-1.42) | | 0.78 (0.47-1.28) | | 0.78 (0.39-1.56) | | 0.96 (0.43-2.11) | |
| Education (highest degree) | | | | | | | | |
| Lower than university | (ref) | 0.377 | (ref) | 0.158 | (ref) | 0.806 | (ref) | **0.001** |
| University | 1.19 (0.81-1.77) | | 0.73 (0.46-1.13) | | 1.08 (0.60-1.93) | | 2.99 (1.54-5.84) | |
| Swiss-SEP[b] | | | | | | | | |
| Per 10-point increase | 1.30 (1.08-1.55) | **0.005** | 1.10 (0.90-1.33) | 0.351 | 1.29 (0.99-1.69) | 0.063 | 1.50 (1.09-2.07) | **0.012** |
| Low | (ref) | 0.077 | (ref) | 0.360 | (ref) | 0.337 | (ref) | 0.140 |
| Medium | 1.44 (0.93-2.22) | | 1.40 (0.88-2.23) | | 1.27 (0.66-2.45) | | 1.64 (0.74-3.64) | |
| High | 1.66 (1.04-2.66) | | 1.21 (0.72-2.04) | | 1.67 (0.84-3.31) | | 2.26 (1.00-5.08) | |
| Marital status | | | | | | | | |

| | | | | | | | | |
|---|---|---|---|---|---|---|---|---|
| Single & Widowed & Divorced | (ref) | 0.060 | (ref) | 0.564 | (ref) | 0.394 | (ref) | **0.003** |
| Married | 0.69 (0.47-1.02) | | 0.89 (0.59-1.34) | | 0.78 (0.44-1.38) | | 0.28 (0.13-0.65) | |
| Children | | | | | | | | |
| No | (ref) | **0.045** | (ref) | 0.221 | (ref) | 0.054 | (ref) | **<0.001** |
| Yes | 0.67 (0.46-0.99) | | 0.77 (0.51-1.17) | | 0.56 (0.31-1.01) | | 0.21 (0.09-0.53) | |
| Living situation | | | | | | | | |
| Alone | (ref) | **0.026** | (ref) | 0.102 | (ref) | 0.259 | (ref) | 0.265 |
| With others | 1.68 (1.06-2.65) | | 1.51 (0.92-46) | | 0.71 (0.39-1.29) | | 1.61 (0.70-3.71) | |
| Smoking status | | | | | | | | |
| Never | (ref) | **<0.001** | (ref) | 0.482 | (ref) | **0.030** | (ref) | **0.002** |
| Former | 1.50 (0.91-2.50) | | 0.81 (0.45-1.44) | | 1.25 (0.59-2.66) | | 1.77 (0.77-4.07) | |
| Current | 4.72 (2.46-9.05) | | 1.40 (0.66-2.96) | | 3.16 (1.34-7.41) | | 3.72 (1.43-9.70) | |
| Physical activity[c] | | | | | | | | |
| Inactive | (ref) | **0.045** | (ref) | 0.621 | (ref) | 0.585 | (ref) | 0.073 |
| Active | 1.57 (1.01-2.47) | | 1.12 (0.71-1.79) | | 1.20 (0.62-2.30) | | 2.33 (0.92-5.85) | |
| SF-36[d] | | | | | | | | |
| Physical functioning | 1.03 (1.01-1.05) | **0.011** | 1.01 (0.99-1.03) | 0.568 | 0.99 (0.97-1.02) | 0.677 | 1.04 (0.99-1.09) | 0.125 |
| Role physical | 1.03 (1.01-1.05) | **0.003** | 1.01 (0.99-1.03) | 0.423 | 1.01 (0.99-1.04) | 0.322 | 1.00 (0.97-1.04) | 0.764 |
| Bodily pain | 1.05 (1.02-1.08) | **0.001** | 1.01 (0.98-1.04) | 0.414 | 1.04 (1.00-1.09) | 0.082 | 1.02 (0.97-1.06) | 0.523 |
| General health | 1.02 (1.00-1.03) | **0.023** | 1.01 (1.00-1.03) | 0.140 | 1.00 (0.98-1.02) | 0.949 | 1.01 (0.98-1.03) | 0.643 |
| Vitality | 1.03 (1.01-1.05) | **0.001** | 1.02 (1.00-1.04) | 0.085 | 1.01 (0.99-1.04) | 0.298 | 1.00 (0.97-1.04) | 0.801 |
| Social functioning | 1.03 (1.01-1.05) | **0.001** | 1.03 (1.01-1.05) | **0.008** | 1.02 (0.99-1.04) | 0.212 | 0.99 (0.97-1.02) | 0.600 |
| Role emotional | 1.02 (1.00-1.04) | 0.086 | 1.02 (1.00-1.04) | 0.138 | 1.00 (0.98-1.03) | 0.871 | 0.98 (0.95-1.00) | 0.078 |
| Mental health | 1.04 (1.01-1.06) | **0.003** | 1.03 (1.01-1.06) | **0.011** | 1.02 (0.98-1.05) | 0.342 | 0.99 (0.95-1.02) | 0.479 |
| Physical component summary | 1.03 (1.00-1.05) | **0.022** | 1.00 (0.98-1.02) | 0.953 | 1.00 (0.97-1.03) | 0.898 | 1.03 (0.99-1.08) | 0.153 |
| Mental component summary | 1.03 (1.01-1.05) | **0.009** | 1.03 (1.00-1.05) | **0.019** | 1.01 (0.98-1.04) | 0.421 | 0.98 (0.95-1.01) | 0.145 |

| | | | | | | | | |
|---|---|---|---|---|---|---|---|---|
| ICCC-3 cancer diagnosis | | | | | | | | |
| I Leukaemias | (ref) | 0.778 | (ref) | 0438 | (ref) | 0.444 | (ref) | 0.347 |
| II Lymphomas | 0.84 (0.51-1.40) | | 0.74 (0.42-1.31) | | 0.64 (0.30-1.35) | | 0.54 (0.21-1.37) | |
| III Central nervous system tumours | 1.01 (0.52-1.99) | | 1.15 (0.56-2.36) | | 0.65 (0.23-1.84) | | 0.52 (0.14-1.98) | |
| Other type | 1.21 (0.76-1.92) | | 1.37 (0.83-2.26) | | 0.79 (0.41-1.55) | | 0.98 (0.46-2.11) | |
| Age at diagnosis, years | | | | | | | | |
| <5 | (ref) | 0.674 | (ref) | 0.539 | (ref) | 0.413 | (ref) | 0.584 |
| 5-9 | 0.84 (0.49-1.44) | | 0.80 (0.46-1.39) | | 0.50 (0.22-1.16) | | 0.69 (0.29-1.67) | |
| 10-14 | 1.17 (0.72-1.91) | | 0.75 (0.44-1.27) | | 0.92 (0.46-1.83) | | 0.61 (0.26-1.42) | |
| ≥15 | 1.01 (0.58-1.74) | | 0.71 (0.39-1.29) | | 0.76 (0.34-1.70) | | 0.57 (0.22-1.52) | |
| Time since diagnosis, years | | | | | | | | |
| ≤25 | (ref) | **0.015** | (ref) | 0.152 | (ref) | 0.798 | (ref) | **0.002** |
| >25 | 0.62 (0.43-0.91) | | 0.74 (0.49-1.12) | | 0.93 (0.53-1.63) | | 0.34 (0.17-0.67) | |
| History of relapse | | | | | | | | |
| No | (ref) | 0.686 | (ref) | 0.211 | (ref) | 0.730 | (ref) | 0.895 |
| Yes | 0.89 (0.52-1.55) | | 0.67 (0.36-1.26) | | 1.15 (0.53-2.48) | | 0.94 (0.36-2.45) | |
| HSCT | | | | | | | | |
| No | (ref) | 0.248 | (ref) | 0.111 | (ref) | 0.115 | (ref) | 0.305 |
| Yes | 2.13 (0.59-7.78) | | 2.89 (0.78-10.68) | | 3.40 (0.74-15.61) | | 2.59 (0.42-15.98) | |
| Cranial radiation | | | | | | | | |
| No | (ref) | 0.580 | (ref) | 0.842 | (ref) | 0.474 | (ref) | 0.474 |
| <18Gy | 0.69 (0.27-1.78) | | 1.26 (0.50-3.17) | | 1.67 (0.52-5.32) | | 1.35 (0.34-5.33) | |
| >=18Gy | 0.80 (0.45-1.43) | | 0.92 (0.49-1.71) | | 1.45 (0.67-3.14) | | 0.49 (0.14-1.72) | |
| Chest radiation | | | | | | | | |
| No | (ref) | 0.496 | (ref) | 0.549 | (ref) | 0.200 | (ref) | 0.844 |
| <30 Gy | 0.69 (0.27-1.78) | | 1.47 (0.60-3.61) | | 0.56 (0.12-2.71) | | 0.91 (0.19-4.47) | |

| | | | | | | | | |
|---|---|---|---|---|---|---|---|---|
| ≥30 Gy | 0.72 (0.37-1.42) | | 0.79 (0.38-1.65) | | 0.28 (0.06-1.25) | | 0.69 (0.19-2.47) | |
| TBI and/or abdominal radiation | | | | | | | | |
| No | (ref) | 0.787 | (ref) | 0.093 | (ref) | 0.261 | (ref) | 0.929 |
| Yes | 1.10 (0.54-2.28) | | 1.85 (0.90-3.81) | | 1.71 (0.67-4.39) | | 0.94 (0.25-3.49) | |
| Anthracyclines | | | | | | | | |
| No | (ref) | 0.285 | (ref) | 0.985 | (ref) | 0.305 | (ref) | 0.084 |
| Yes | 1.24 (0.84-1.83) | | 1.00 (0.65-1.52) | | 1.35 (0.76-2.37) | | 1.79 (0.93-3.45) | |
| Glucocorticoids | | | | | | | | |
| No | (ref) | 0.613 | (ref) | 0.098 | (ref) | 0.711 | (ref) | 0.544 |
| Yes | 0.91 (0.62-1.33) | | 0.70 (0.46-1.07) | | 1.11 (0.63-1.97) | | 0.81 (0.42-1.58) | |
| Alkylating agents | | | | | | | | |
| No | (ref) | 0.578 | (ref) | 0.472 | (ref) | 0.834 | (ref) | 0.554 |
| Yes | 1.11 (0.76-1.62) | | 0.86 (0.57-1.30) | | 1.06 (0.61-1.86) | | 1.22 (0.63-2.34) | |

*Abbreviations: CI, confidence intervals; Gy, Gray; HSCT, haematopoietic stem cell transplantation; ICCC-3, International Classification of Childhood Cancer, Third Edition; OR, odds ratios; SEP, socioeconomic position; SF-36, 36-Item Short Form Survey; TBI, total body irradiation.*

[a] *p* - value was calculated using Wald test.

[b] Swiss neighbourhood index of socioeconomic position (Swiss-SEP), a validated area-based index (range 0–100) reflecting household education and occupation, housing conditions, and rent values. For descriptive analyses, values were categorised into tertiles (low, medium, high) based on the national Swiss population distribution. In regression models, Swiss-SEP was analyzed both as a continuous variable (odds ratio per 10-point increase) and as tertiles.

[c] PA active: ≥150 min of moderate intense or 75 min of vigorous intense or a combination of moderate and vigorous intense physical activity per week.

[d] SF-36, all scores are norm-based (mean = 50, SD = 10). Higher scores indicate better health-related quality of life. The physical component summary score is primarily derived from physical functioning, role physical, bodily pain, and general health domains, whereas the mental component summary score is primarily derived from vitality, social functioning, role emotional, and mental health domains.

Supplementary Table 6. Association of sociodemographic, lifestyle and clinical characteristics by CVD risk profiles (univariable multinomial logistic regressions).

| | Compared to CVD risk-free[a] | | | |
|---|---|---|---|---|
| | CVD[b] | | CVD risk factors[c] | |
| | OR (95% CI) | *p* - value[d] | OR (95% CI) | *p* - value[d] |
| Sex | | | | |
| Female | (ref) | 0.218 | (ref) | 0.245 |
| Male | 0.78 (0.52-1.16) | | 1.31 (0.83-2.05) | |
| Age at survey, years | | | | |
| ≤30 | (ref) | **<0.001** | (ref) | **<0.001** |
| 31-39 | 1.10 (0.65-1.86) | | 1.57 (0.84-2.96) | |
| ≥40 | 2.41 (1.44-4.03) | | 3.61 (1.95-6.67) | |
| Language region within Switzerland | | | | |
| German speaking | (ref) | 0.922 | (ref) | 0.914 |
| French speaking | 0.96 (0.61-1.51) | | 0.90 (0.54-1.51) | |
| Italian speaking | 0.74 (0.16-3.31) | | 0.95 (0.21-4.26) | |
| Migration background | | | | |
| Swiss, no migration background | (ref) | 0.097 | (ref) | 0.374 |
| Migration background | 0.62 (0.35-1.09) | | 1.27 (0.75-2.13) | |
| Education (highest degree) | | | | |
| Lower than university | (ref) | 0.410 | (ref) | **<0.001** |
| University | 0.84 (0.55-1.28) | | 0.33 (0.18-0.60) | |
| Swiss-SEP[e] | | | | |
| Per 10-point increase | 1.08 (0.89-1.32) | 0.409 | 0.98 (0.79-1.21) | 0.846 |
| Low | (ref) | 0.626 | (ref) | 0.358 |
| Medium | 1.03 (0.64-1.66) | | 0.68 (0.40-1.16) | |
| High | 1.26 (0.76-2.07) | | 0.88 (0.51-1.52) | |
| Marital status | | | | |
| Single & Widowed & Divorced | (ref) | 0.341 | (ref) | 0.738 |
| Married | 1.22 (0.81-1.84) | | 1.08 (0.68-1.72) | |
| Children | | | | |
| No | (ref) | 0.248 | (ref) | 0.665 |
| Yes | 1.28 (0.84-1.93) | | 0.90 (0.55-1.46) | |
| Living situation | | | | |
| Alone | (ref) | 0.734 | (ref) | 0.247 |
| With others | 0.92 (0.56-1.50) | | 0.73 (0.44-1.24) | |
| Smoking status | | | | |
| Never | (ref) | 0.361 | (ref) | 0.646 |
| Former | 1.33 (0.80-2.22) | | 0.73 (0.37-1.43) | |

| | | | | |
|---|---|---|---|---|
| Current | 0.82 (0.45-1.48) | | 0.91 (0.49-1.68) | |
| Physical activity[f] | | | | |
| Inactive | (ref) | 0.331 | (ref) | 0.631 |
| Active | 0.79 (0.49-1.27) | | 0.88 (0.51-1.50) | |
| SF-36[g] | | | | |
| Physical functioning | 0.96 (0.94-0.98) | **<0.001** | 0.97 (0.95-0.99) | **0.015** |
| Role physical | 0.96 (0.94-0.98) | **<0.001** | 0.98 (0.96-1.01) | 0.153 |
| Bodily pain | 0.94 (0.92-0.97) | **<0.001** | 0.97 (0.94-1.01) | 0.117 |
| General health | 0.96 (0.94-0.97) | **<0.001** | 0.98 (0.96-1.00) | **<0.039** |
| Vitality | 0.97 (0.95-0.99) | **0.001** | 0.98 (0.96-1.00) | **0.049** |
| Social functioning | 0.98 (0.96-0.99) | **0.009** | 0.98 (0.96-1.00) | **0.033** |
| Role emotional | 0.98 (0.96-0.99) | **0.007** | 0.99 (0.97-1.01) | 0.422 |
| Mental health | 0.98 (0.96-1.00) | 0.095 | 0.97 (0.95-1.00) | **0.042** |
| Physical component summary | 0.94 (0.93-0.96) | **<0.001** | 0.97 (0.95-1.00) | **0.041** |
| Mental component summary | 0.98 (0.96-1.00) | 0.050 | 0.98 (0.96-1.00) | 0.075 |
| ICCC-3 cancer diagnosis | | | | |
| I Leukaemias | (ref) | 0.561 | (ref) | 0.105 |
| II Lymphomas | 1.00 (0.57-1.76) | | 0.88 (0.46-1.69) | |
| III Central nervous system tumours | 0.63 (0.27-1.50) | | 1.87 (0.95-3.73) | |
| Other type | 1.09 (0.67-1.76) | | 0.84 (0.48-1.48) | |
| Age at diagnosis, years | | | | |
| <5 | (ref) | 0.119 | (ref) | 0.169 |
| 5-9 | 1.59 (0.89-2.83) | | 1.56 (0.83-2.94) | |
| 10-14 | 1.24 (0.71-2.14) | | 1.41 (0.79-2.52) | |
| ≥15 | 1.99 (1.13-3.52) | | 1.25 (0.63-2.51) | |
| Time since diagnosis, years | | | | |
| ≤25 | (ref) | **0.004** | (ref) | **0.013** |
| >25 | 1.83 (1.21-2.77) | | 1.80 (1.13-2.86) | |
| History of relapse | | | | |
| No | (ref) | 0.545 | (ref) | 0.893 |
| Yes | 1.20 (0.67-2.14) | | 1.05 (0.53-2.06) | |
| HSCT | | | | |
| No | (ref) | 0.369 | (ref) | 0.965 |
| Yes | 1.53 (0.60-3.88) | | 0.97 (0.28-3.33) | |
| Cranial radiation | | | | |
| No | (ref) | 0.728 | (ref) | **0.006** |
| <18 Gy | 0.79 (0.27-2.31) | | 2.10 (0.88-5.04) | |
| ≥18 Gy | 1.22 (0.66-2.28) | | 2.39 (1.33-4.31) | |
| Chest radiation | | | | |
| No | (ref) | 0.628 | (ref) | 0.634 |

| | | | | |
|---|---|---|---|---|
| <30 Gy | 1.51 (0.64-3.60) | | 1.06 (0.36-3.13) | |
| ≥30 Gy | 1.11 (0.52-2.34) | | 0.60 (0.21-1.72) | |
| TBI and/or abdominal radiation | | | | |
| No | (ref) | 0.648 | (ref) | 0.157 |
| Yes | 1.17 (0.60-2.25) | | 0.47 (0.17-1.34) | |
| Anthracyclines | | | | |
| No | (ref) | 0.636 | (ref) | 0.053 |
| Yes | 1.10 (0.73-1.66) | | 0.61 (0.37-1.01) | |
| Glucocorticoids | | | | |
| No | (ref) | 0.291 | (ref) | 0.196 |
| Yes | 0.79 (0.51-1.22) | | 1.25 (0.89-1.77) | |
| Alkylating agents | | | | |
| No | (ref) | 0.734 | (ref) | 0.267 |
| Yes | 1.07 (0.71-1.62) | | 0.82 (0.58-1.16) | |

*Abbreviations: CI, confidence intervals; Gy, Gray; HSCT, haematopoietic stem cell transplantation; ICCC-3, International Classification of Childhood Cancer, Third Edition; OR, odds ratios; SEP, socioeconomic position; SF-36, 36-Item Short Form Survey; TBI, total body irradiation.*

[a] "CVD risk-free" if survivors did not report any CVD conditions and CVD risk factors.

[b] "CVD" including atrial fibrillation, deep venous thrombosis, cardiomyopathy, stroke/transient ischemic attack, angina pectoris, heart attack, arteriosclerosis.

[c] "CVD risk factors" including hypertension (repeated high blood pressure measurements or antihypertensive medication treatment), obesity, diabetes mellitus treated with either tablets or insulin, and/or high cholesterol defined as treatment with lipid-lowering medications.

[d] *p* - value was calculated using Wald test.

[e] Swiss neighbourhood index of socioeconomic position (Swiss-SEP), a validated area-based index (range 0–100) reflecting household education and occupation, housing conditions, and rent values. For descriptive analyses, values were categorised into tertiles (low, medium, high) based on the national Swiss population distribution. In regression models, Swiss-SEP was analysed both as a continuous variable (odds ratio per 10-point increase) and as tertiles.

[f] PA active: ≥150 min of moderate intense or 75 min of vigorous intense or a combination of moderate and vigorous intense physical activity per week.

[g] SF-36, all scores are norm-based (mean = 50, SD = 10). Higher scores indicate better health-related quality of life. The physical component summary score is primarily derived from physical functioning, role physical, bodily pain, and general health domains, whereas the mental component summary score is primarily derived from vitality, social functioning, role emotional, and mental health domains.
